# Deep learning-enabled MRI phenotyping uncovers regional body composition heterogeneity and disease associations in two European population cohorts

**DOI:** 10.1101/2025.06.03.25328867

**Authors:** Christian J. Mertens, Hartmut Häntze, Sebastian Ziegelmayer, Jakob Nikolas Kather, Daniel Truhn, Su Hwan Kim, Felix Busch, Dominik Weller, Benedikt Wiestler, Markus Graf, Fabian Bamberg, Christopher L. Schlett, Jakob B. Weiss, Steffen Ringhof, Elif Can, Jeanette Schulz-Menger, Thoralf Niendorf, Jacqueline Lammert, Isabel Molwitz, Avan Kader, Alessa Hering, Aymen Meddeb, Jawed Nawabi, Matthias B. Schulze, Thomas Keil, Stefan N. Willich, Lilian Krist, Martin Hadamitzky, Anke Hannemann, Florian Bassermann, Daniel Rueckert, Tobias Pischon, Alexander Hapfelmeier, Marcus R. Makowski, Keno K. Bressem, Lisa C. Adams

**Author notes:** contributed equally as first authors (Christian J. Mertens, Hartmut Häntze). contributed equally as senior authors (Keno K. Bressem, Lisa C. Adams). Corresponding authors: Christian Mertens, MD, and Keno Bressem, MD.

## Abstract

Body mass index (BMI) does not account for substantial inter-individual differences in regional fat and muscle compartments, which are relevant for the prevalence of cardiometabolic and cancer conditions. We applied a validated deep learning pipeline for automated segmentation of whole-body MRI scans in 45,851 adults from the UK Biobank and German National Cohort, enabling harmonized quantification of visceral (VAT), gluteofemoral (GFAT), and abdominal subcutaneous adipose tissue (ASAT), liver fat fraction (LFF), and trunk muscle volume. Associations with clinical conditions were evaluated using compartment measures adjusted for age, sex, height, and BMI. Our analysis demonstrates that regional adiposity and muscle volume show distinct associations with cardiometabolic and cancer prevalence, and that substantial disease heterogeneity exists within BMI strata. The analytic framework and reference data presented here will support future risk stratification efforts and facilitate the integration of automated MRI phenotyping into large-scale population and clinical research.

## Introduction

Obesity affects nearly 890 million people worldwide and represents a major public health challenge with far-reaching clinical implications. While substantial evidence has established obesity as a potent driver of cardiovascular and metabolic disease,^1–3^ an expanding body of research now also suggests links to various malignancies including colorectal, pancreatic, prostate, and breast cancer, albeit with variable effect magnitudes.^4–9^

Body mass index (BMI) remains the most widely used clinical tool worldwide for obesity assessment, as it is simple to calculate and implement. However, BMI exhibits notable limitations in its inability to adequately reflect the distribution and metabolic characteristics of adipose tissue, factors increasingly recognized as crucial correlates of health conditions beyond overall body weight.^10,11^ The heterogeneity in metabolic phenotypes across BMI categories further underscores these limitations. Individuals classified as obese by BMI criteria but exhibiting preserved metabolic function and physical fitness, often termed metabolically healthy obese (MHO),^11^ frequently present with more favourable cardiometabolic profiles than their BMI would suggest.^11–14^ Conversely, individuals with normal BMI but disproportionate accumulation of visceral adipose tissue, described as metabolically obese normal weight (MONW), exhibit elevated risk of metabolic dysfunction and cardiovascular disease.^15^ These contrasting phenotypes underscore metabolic heterogeneity, suggesting that regional body composition, rather than overall weight, may better reflect variation in the prevalence of cardiometabolic, cancer, and related clinical conditions.

Still, the precise quantification of body composition presents with methodological challenges. While anthropometric indices such as waist to hip ratio provide incremental information beyond BMI, magnetic resonance imaging (MRI) enables a highly accurate non-invasive assessment of adipose tissue compartments. MRI allows for detailed quantification of visceral adipose tissue (VAT), gluteofemoral adipose tissue (GFAT), abdominal subcutaneous adipose tissue (ASAT), trunk muscle volume, and liver fat fraction (LFF). Manual delineation, however, remains labour-intensive and has historically limited studies to a few hundred scans from a single imaging protocol.

Deep-learning approaches allow us to overcome that bottleneck and enable fully automated extraction of regional body composition metrics from tens of thousands of MRIs.^16–18^ We analysed whole body MRIs from 45,851 participants from UK Biobank (UKB)^19^ and the German National Cohort (NAKO).^20^ ^21^ These two population studies were scanned on different hardware, providing a natural test bed for cross-platform harmonisation.^22,23^

Using deep learning-based segmentation techniques, we quantified VAT, GFAT, ASAT, muscle and LFF in each participant and examined their independent associations with nine prevalent diseases spanning cardiometabolic, cancer and musculoskeletal domains. For each body composition compartment, associations with clinical conditions were estimated after adjusting for age, sex, height, and BMI. Odds ratios are reported per 1 SD higher compartment volume at a given BMI and body size, reflecting redistribution of body composition rather than absolute increases. By integrating data from two population-based cohorts scanned with different MRI protocols, we tested whether regional body composition phenotypes provide generalisable and BMI-independent associations with prevalence of cardiometabolic, cancer, and related clinical conditions, reflecting variation in fat and muscle distribution at a given BMI and body size rather than absolute compartment volumes.

## Results

### Cohort characteristics and prevalence of cardiometabolic, cancer, and related clinical conditions

We analysed whole-body MRIs in 45,851 adults from two large imaging cohorts: 26,877 of the first 30,861 NAKO participants and 18,974 of the first 19,512 UK Biobank (UKB) participants. The NAKO subsample included 14,969 men (55.7%) and 11,908 women (44.3%), consistent with the full imaging dataset. The UKB imaging cohort was sex-balanced (9,094 men, 47.9%; 9,880 women, 52.1%) but older, due to targeted recruitment of participants aged ≥55 years at the outset of imaging. Median age was 63 years (IQR 56–68) in UKB versus 48 years (IQR 40–58) in NAKO (p□<□0.0001; Fig. 1, Supplementary Tables S1–S2). Clinical conditions analysed include diseases (e.g., CAD, diabetes, cancer) as well as risk factors and symptoms (e.g., hyperlipidaemia, osteoporosis, gout, back pain).

**Figure 1.**
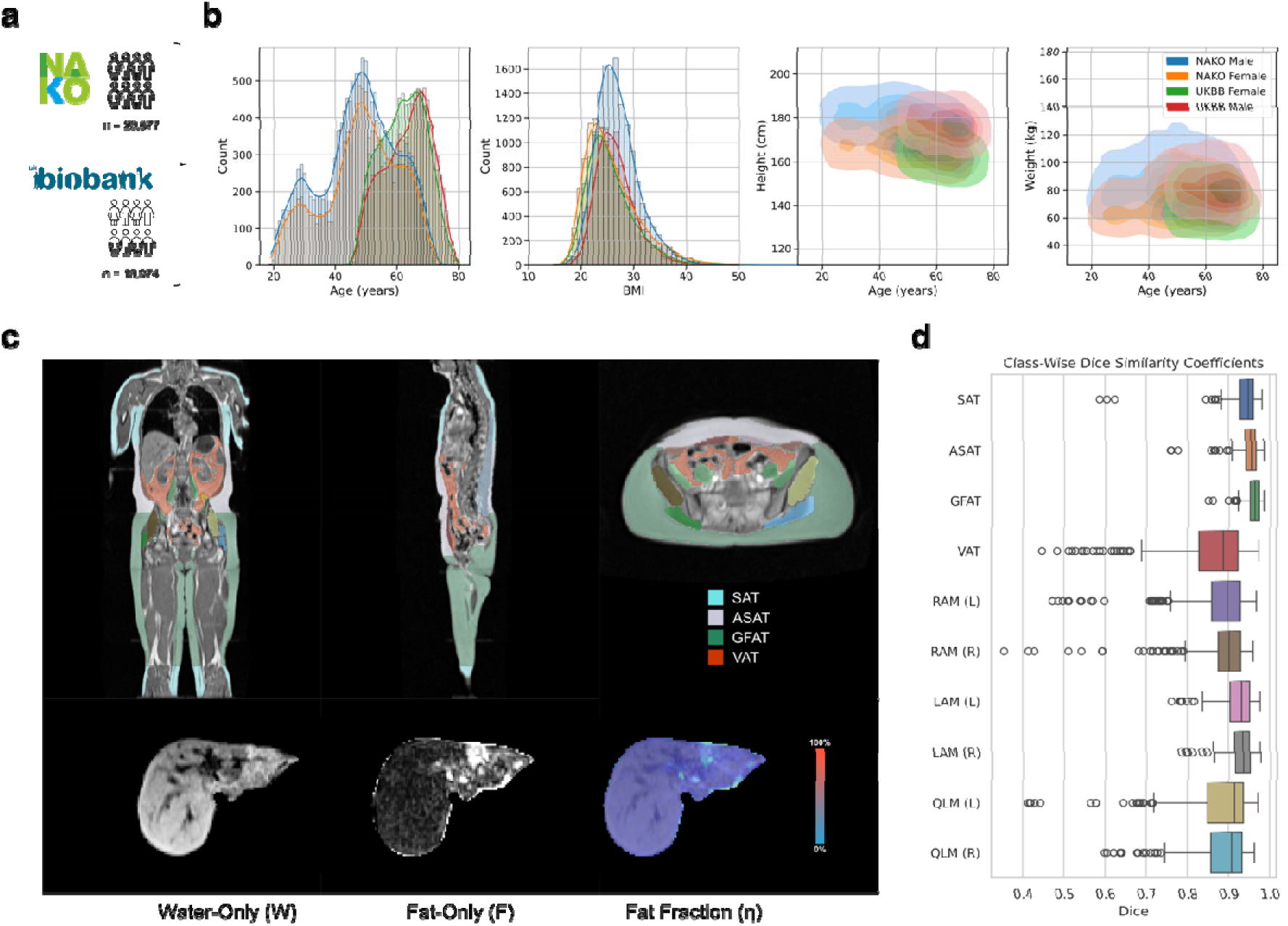
Deep learning-based segmentation of body composition from MRI scans. (**a**) A total of 26,877 NAKO and 18,974 UKB participants were included based on available imaging, lifestyle variables, and outcome data. (**b**) Histograms and density plots of participants baseline characteristics stratified by dataset and sex. (**c**) Representative segmentation results illustrating subcutaneous adipose tissue (SAT), abdominal subcutaneous adipose tissue (ASAT), gluteofemoral adipose tissue (GFAT), visceral adipose tissue (VAT), and trunk musculature, including rectus abdominis muscles (RAM), lateral abdominal muscles (LAM), and quadratus lumborum muscles (QLM). Segmented liver images are shown, including water-only, fat-only, and fat fraction maps. Fat fraction was calculated via the ratio of fat signal intensity over combined signal intensity: η = F/(W+F). (**d**) Dice similarity coefficients (DSC) for each segmented tissue class, evaluated using 5-fold participant-stratified cross-validation. Box plots display the median and interquartile range, with whiskers extending to the minimum and maximum values within 1.5 times the IQR, and outliers shown as individual points. The model exhibited high segmentation accuracy across most compartments, with DSC values of 0.94 for SAT, 0.95 for ASAT, and 0.96 for GFAT (UKB data only as GFAT is not included in NAKO T2-HASTE training sequences). VAT segmentation showed the lowest DSC (0.85) likely due to its higher spatial complexity. Muscle segmentation was consistent between sides, with DSC values of 0.92/0.93 for LAM, and 0.87/0.88 for both RAM and QLM (left/right).

BMI distributions were similar between cohorts (men: 26.3□kg/m² in NAKO, 26.0□kg/m² in UKB; women: 24.8 and 24.7□kg/m², respectively), but prevalence of cardiometabolic conditions varied considerably (Table□1). Coronary artery disease (CAD) was 5.5% in UKB men and 1.2% in NAKO men, a 4.6-fold difference. This likely reflects both age structure and differing definitions: NAKO relied on self-reported myocardial infarction, angina, or coronary stenosis; UKB used a combination of hospital records, self-report, and procedure codes. T2D prevalence was higher in UKB men (7.0% vs. 5.2%) but slightly lower in women (3.5% vs. 4.4%). Hyperlipidaemia was also more frequent in UKB (men: 52.3% vs. 46.3%; women: 45.3% vs. 37.0%). In contrast, heart failure was reported more often in NAKO (1.3%) than in UKB (0.1%), likely reflecting differences in questionnaire structure, self-report practices, or diagnostic coding.

**Table 1:** Characteristics of NAKO and UKB participants.

| Dataset | NAKO |  | UKB |  |
| --- | --- | --- | --- | --- |
| Sex | Male<br>(n = 14,969) | Female<br>(n = 11,908) | Male<br>(n = 9,094) | Female<br>(n = 9,880) |
| Age (years) | 48 [40, 57] | 49 [41, 58] | 64 [58, 69] | 62 [56, 68] |
| Ethnicity |  |  |  |  |
| White | 14,148<br>(94.51%) | 11,050 (92.79%) | 8,779 (96.54%) | 9601<br>(97.18%) |
| Not specified | 663 (4.43%) | 730 (6.13%) |  |  |
| Other* | 160 (1.07%) | 128 (1.07%) | 315 (3.46%) | 279 (2.82%) |
| Weight (kg) | 85 [76, 94] | 68 [61, 78] | 82 [75, 90] | 66 [60, 75] |
| Height (cm) | 179 [174, 184] | 166 [161, 170] | 177 [173, 182] | 163 [159, 168] |
| Body mass index (kg/m<sup>2</sup>) | 26.3 [24.1, 29.1] | 24.8 [22.1, 28.6] | 26 [23.9, 28.5] | 24.7 [22.4, 27.9] |
| Hand grip strength (kg) | 48.7 [43.2, 54.7] | 30.4 [26.7, 34.2] |  |  |
| Physical activity (MET min. / week) |  |  |  |  |
| Insufficient (< 600) | 2047 (13.67%) | 1751 (14.7%) |  |  |
| Sufficient (600 – 999) | 931 (6.22%) | 797 (6.69%) |  |  |
| Moderate (1000 – 2999) | 3770 (25.19%) | 2954 (24.81%) |  |  |
| High (≥ 3000) | 8221 (54.92%) | 6406 (53.8%) |  |  |
| Alcohol |  |  |  |  |
| Never |  |  | 489 (5.38%) | 720 (7.29%) |
| Special occasions only |  |  | 589 (6.48%) | 1359<br>(13.76%) |
| One to three times a month |  |  | 864 (9.5%) | 1361<br>(13.78%) |
| Once or twice a week |  |  | 2373 (26.09%) | 2736<br>(27.69%) |
| Three or four times a week |  |  | 2903 (31.92%) | 2413<br>(24.42%) |
| Daily or almost daily |  |  | 1876 (20.63%) | 1291<br>(13.07%) |
| Smoking |  |  |  |  |
| Never |  |  | 5265 (57.9%) | 6532<br>(66.11%) |
| Previous |  |  | 3414 (37.54%) | 3021<br>(30.58%) |
| Current |  |  | 415 (4.56%) | 327 (3.31%) |
| **Body composition metrics** |  |  |  |  |
| Subcutaneous adipose tissue (L) | 12.67 [9.58, 16.43] | 17.79 [13.42, 23.49] | 11.31 [8.65, 14.64] | 17.79 [13.42, 23.49] |
| Abdominal subcutaneous adipose tissue (L) | 3.98 [2.78, 5.55] | 4.50 [2.99, 6.63] | 3.47 [2.54, 4.77] | 4.44 [3.11, 6.26] |
| Gluteofemoral adipose tissue (L) | 4.82 [3.69, 6.26] | 8.69 [6.81, 11.12] | 4.16 [3.13, 5.55] | 7.75 [5.93, 10.06] |
| Visceral adipose tissue (L) | 2.69 [1.43, 4.14] | 1.06 [0.59, 2.00] | 3.10 [2.03, 4.38] | 1.47 [0.98, 2.28] |
| Total trunk muscle volume (L) | 6.38 [5.82, 7.00] | 4.74 [4.34, 5.21] | 6.21 [5.57, 6.91] | 4.40 [3.97, 4.87] |
| Liver fat fraction (%) | 7.45 [5.95, 10.87] | 5.75 [4.71, 7.64] | 8.44 [7.29, 10.65] | 7.44 [6.52, 8.89] |
| **Disease outcomes** |  |  |  |  |
| **Cardiovascular** |  |  |  |  |
| Hypertension | 3846 (25.69%) | 2517 (21.14%) | 2618 (28.79%) | 1888<br>(19.11%) |
| CAD | 180 (1.20%) | 70 (0.59%) | 501 (5.51%) | 166 (1.68%) |
| Heart failure | 189 (1.26%) | 158 (1.33%) | 9 (0.1%) | 9 (0.09%) |
| **Metabolic** |  |  |  |  |
| Type 2 diabetes | 773 (5.16%) | 529 (4.44%) | 639 (7.03%) | 349 (3.53%) |
| Hyperlipidaemia | 6937 (46.34%) | 4408 (37.02%) | 4752 (52.25%) | 4473<br>(45.27%) |
| Gout | 781 (5.22%) | 199 (1.67%) | 270 (2.97%) | 18 (0.18%) |
| **Other** |  |  |  |  |
| Cancer |  |  | 972 (10.69%) | 1175 (11.89%) |
| Back pain | 2854 (19.07%) | 2837 (23.82%) | 192 (2.11%) | 187 (1.89%) |
| Osteoporosis | 161 (1.08%) | 403 (3.38%) | 26 (0.29%) | 285 (2.88%) |

Note: Continuous variables are shown as median with interquartile range in square brackets. Number of participants corresponding to a specific group are shown with relative abundance in percent.
\* Other ethnicities specified in Supplementary Tables S1 and S2

### Age and sex differences in body composition

We observed consistent age- and sex-related differences in body composition across both cohorts. VAT increased markedly with age, particularly in men (Figure 2a), with VAT volumes 179% higher in women and 236% higher in men at age 60–70 compared to 20–30 years (Table 3). In contrast, GFAT declined with age, decreasing by 5% in women and 9% in men over the same age range (Supplementary Figure S1).

**Figure 2.**
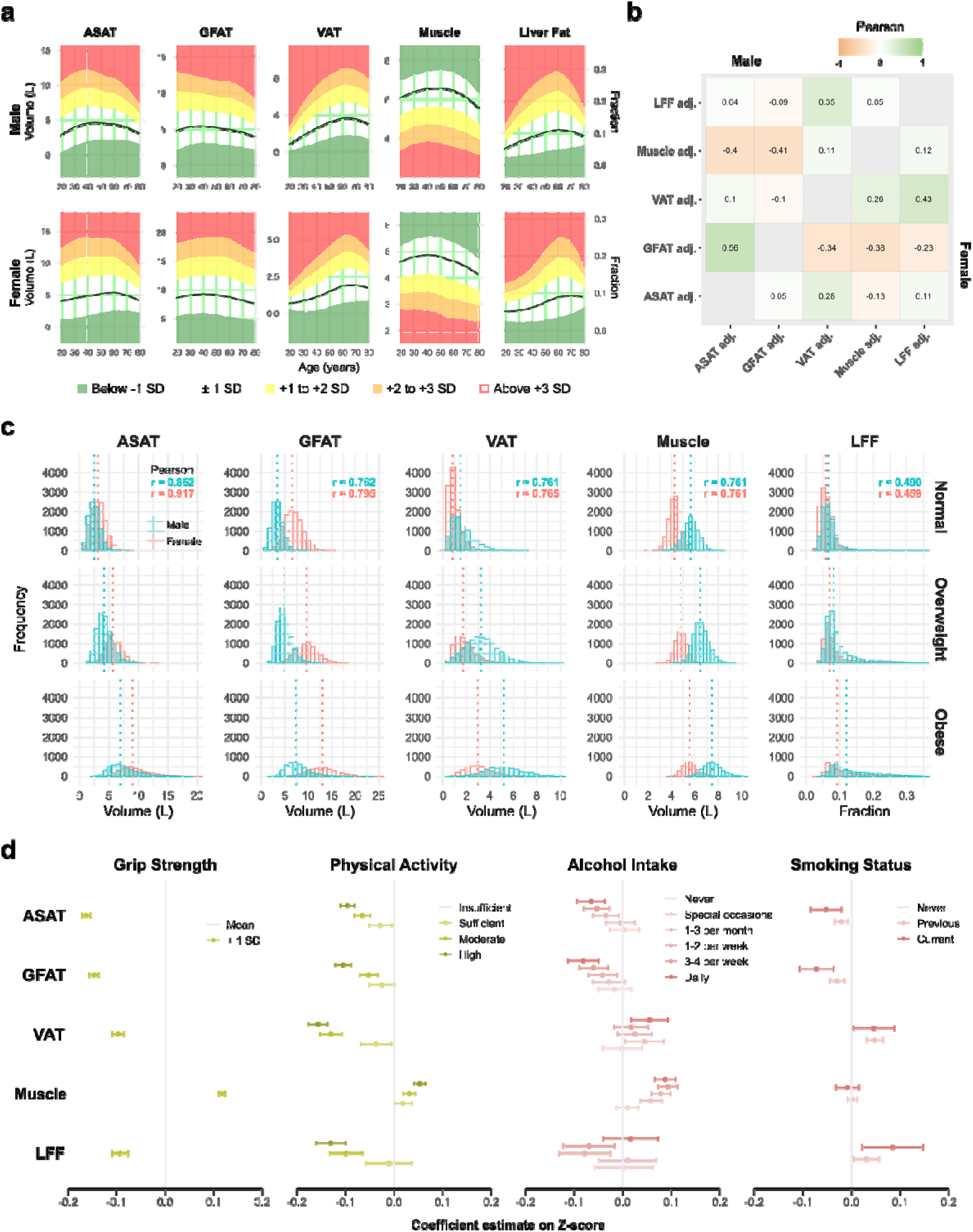
Body composition parameters and associations with age, BMI, and health behaviour indicators. (a) Age-related patterns in body composition parameters (BCPs), shown as LOESS curves with cohort-specific means (black lines) and ±1 standard deviation (shaded areas). (b) Correlation matrix of adjusted BCPs (residuals after adjustment for age, height, and BMI), with Pearson coefficients presented for men (upper triangle) and women (lower triangle); all correlations p_J<_J0.0001 from two-sided t-tests. (c) Distributions of BCPs by BMI category for men and women, displayed as histograms with dashed lines indicating medians. Pearson correlation coefficients between each BCP and BMI are shown in the top row. (d) Estimated associations between health behaviour indicators and unadjusted BCP Z-scores, derived from linear mixed-effects models stratified by cohort and adjusted for age, height, BMI, sex (if applicable), and available behavioural covariates (hand grip strength and physical activity in NAKO; smoking and alcohol intake in UKB). Imaging centre was included as a random intercept. All associations reflect cross-sectional patterns and are not indicative of causal relationships.

**Table 2:** Dice Similarity Coefficients (DSC) for all classes. We used 5-fold participant-stratified cross validation on UKB Dixon in-phase, opposed-phase, water-only and fat-only (in, opp, W, F) as well as NAKO T2-HASTE data (T2). Classes are subcutaneous adipose tissue (SAT), abdominal subcutaneous adipose tissue (ASAT), gluteofemoral adipose tissue (GFAT), visceral adipose tissue (VAT), left and right rectus abdominis muscle (RAM), left and right lateral abdominal muscles (LAM), left and right quadratus lumborum muscle (QLM). GFAT is not included in the NAKO T2-HASTE sequences, so the values are for the UKB data only. The table shows the mean DSC ± standard deviation.

|  | SAT | ASAT | GFAT | VAT | RAM<br>(L) | RAM<br>(R) | LAM<br>(L) | LAM<br>(R) | QLM<br>(L) | QLM<br>(R) | Average |
| --- | --- | --- | --- | --- | --- | --- | --- | --- | --- | --- | --- |
| **F** | 0.93 ± 0.05 | 0.95 ± 0.03 | 0.96 ± 0.02 | 0.85 ± 0.11 | 0.86 ± 0.10 | 0.88 ± 0.09 | 0.92 ± 0.04 | 0.93 ± 0.03 | 0.86 ± 0.10 | 0.88 ± 0.07 | **0.90 ± 0.07** |
| **T2** | 0.96 ± 0.02 | 0.96 ± 0.03 | - | 0.90 ± 0.05 | 0.91 ± 0.04 | 0.91 ± 0.05 | 0.95 ± 0.02 | 0.95 ± 0.03 | 0.92 ± 0.09 | 0.92 ± 0.06 | **0.93 ± 0.04** |
| **W** | 0.92 ± 0.05 | 0.94 ± 0.03 | 0.95 ± 0.02 | 0.81 ± 0.11 | 0.85 ± 0.10 | 0.86 ± 0.10 | 0.91 ± 0.04 | 0.92 ± 0.03 | 0.85 ± 0.11 | 0.87 ± 0.08 | **0.89 ± 0.07** |
| **in** | 0.95 ± 0.02 | 0.95 ± 0.02 | 0.97 ± 0.01 | 0.89 ± 0.06 | 0.89 ± 0.09 | 0.90 ± 0.08 | 0.93 ± 0.03 | 0.94 ± 0.03 | 0.87 ± 0.10 | 0.89 ± 0.07 | **0.92 ± 0.05** |
| **opp** | 0.93 ± 0.05 | 0.94 ± 0.03 | 0.96 ± 0.02 | 0.81 ± 0.13 | 0.84 ± 0.10 | 0.86 ± 0.10 | 0.91 ± 0.04 | 0.91 ± 0.03 | 0.85 ± 0.10 | 0.87 ± 0.08 | **0.89 ± 0.07** |
| **Average** | **0.94 ± 0.04** | **0.95 ± 0.03** | **0.96 ± 0.02** | **0.85 ± 0.10** | **0.87 ± 0.09** | **0.88 ± 0.09** | **0.92 ± 0.04** | **0.93 ± 0.03** | **0.87 ± 0.10** | **0.88 ± 0.07** | **0.91 ± 0.06** |

**Table 3:**
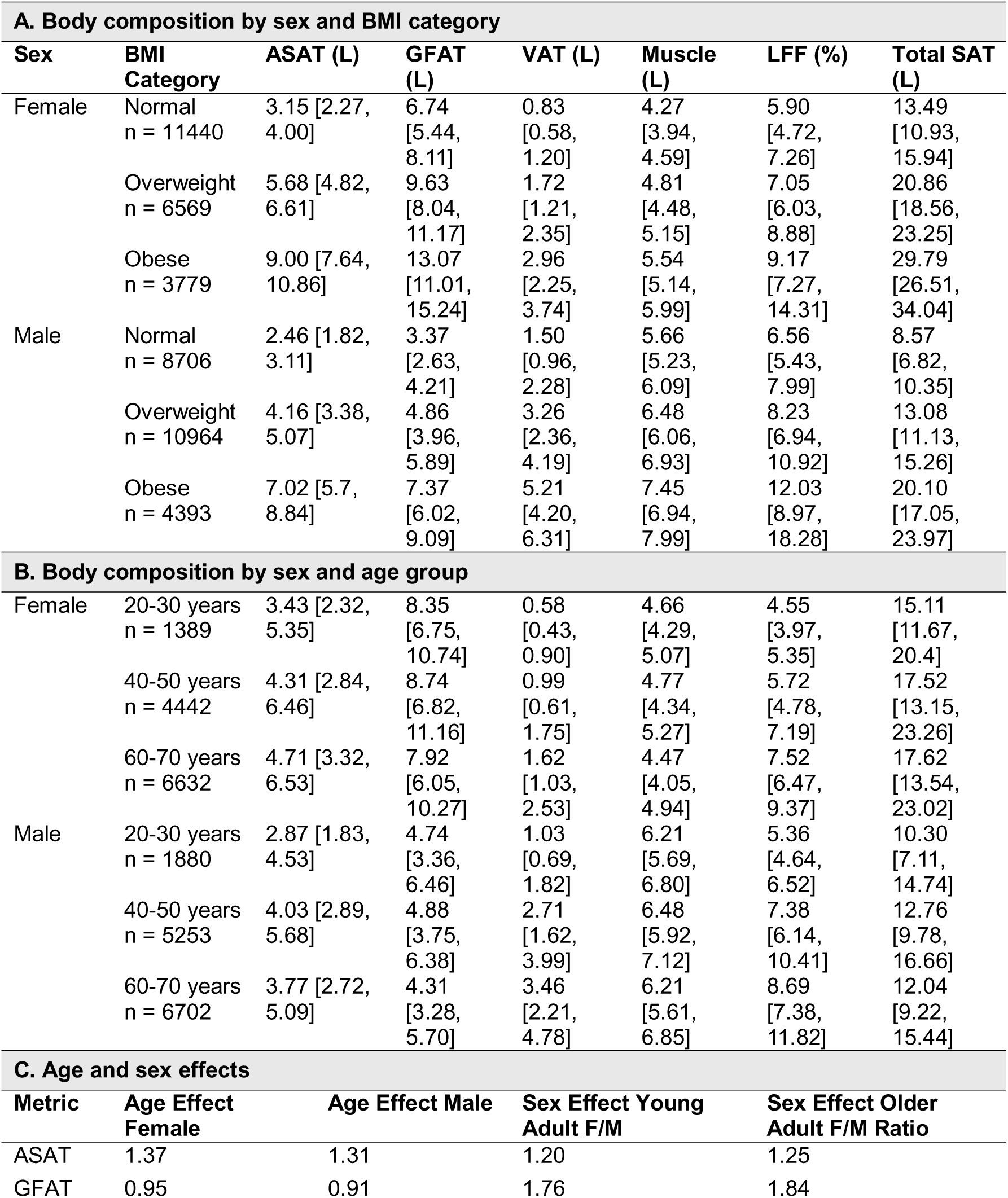
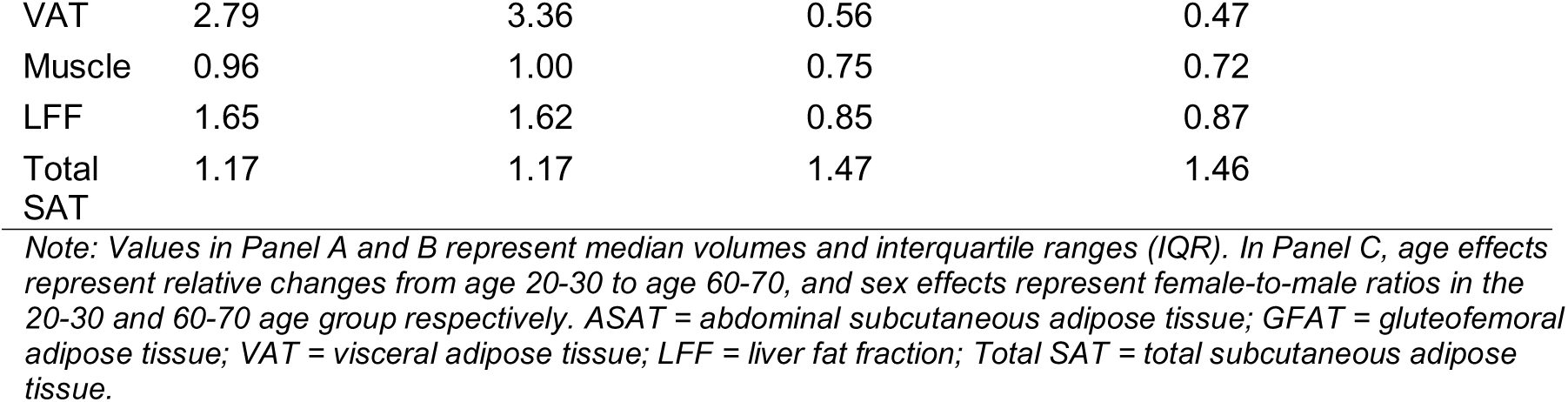
Body composition distribution by sex, age, and BMI category. Table 3 integrates both BMI and age effects on body composition, providing a complete overview of the primary determinants of fat and muscle distribution. The table highlights how body fat compartments differ markedly between sexes, change systematically with aging, and vary across BMI categories. Panel A presents pooled median values and interquartile ranges (IQR) categorized by sex and BMI category based on harmonized data from the UK Biobank (UKB) and German National Cohort (NAKO). Panel B presents median and IQR by sex and age group. Panel C summarizes relative age-related changes and sex-specific ratios, showing percentage changes from early to late adulthood (20–30 years to 60–70 years) and female-to-male ratios in early and late adulthood.

| **A. Body composition by sex and BMI category** |  |  |  |  |  |  |  |
| --- | --- | --- | --- | --- | --- | --- | --- |
| **Sex** | **BMI Category** | **ASAT (L)** | **GFAT (L)** | **VAT (L)** | **Muscle (L)** | **LFF (%)** | **Total SAT (L)** |
| Female | Normal<br>n = 11440 | 3.15 [2.27, 4.00] | 6.74 [5.44, 8.11] | 0.83 [0.58, 1.20] | 4.27 [3.94, 4.59] | 5.90 [4.72, 7.26] | 13.49 [10.93, 15.94] |
|  | Overweight<br>n = 6569 | 5.68 [4.82, 6.61] | 9.63 [8.04, 11.17] | 1.72 [1.21, 2.35] | 4.81 [4.48, 5.15] | 7.05 [6.03, 8.88] | 20.86 [18.56, 23.25] |
|  | Obese<br>n = 3779 | 9.00 [7.64, 10.86] | 13.07 [11.01, 15.24] | 2.96 [2.25, 3.74] | 5.54 [5.14, 5.99] | 9.17 [7.27, 14.31] | 29.79 [26.51, 34.04] |
| Male | Normal<br>n = 8706 | 2.46 [1.82, 3.11] | 3.37 [2.63, 4.21] | 1.50 [0.96, 2.28] | 5.66 [5.23, 6.09] | 6.56 [5.43, 7.99] | 8.57 [6.82, 10.35] |
|  | Overweight<br>n = 10964 | 4.16 [3.38, 5.07] | 4.86 [3.96, 5.89] | 3.26 [2.36, 4.19] | 6.48 [6.06, 6.93] | 8.23 [6.94, 10.92] | 13.08 [11.13, 15.26] |
|  | Obese<br>n = 4393 | 7.02 [5.7, 8.84] | 7.37 [6.02, 9.09] | 5.21 [4.20, 6.31] | 7.45 [6.94, 7.99] | 12.03 [8.97, 18.28] | 20.10 [17.05, 23.97] |
| **B. Body composition by sex and age group** |  |  |  |  |  |  |  |
| Female | 20-30 years<br>n = 1389 | 3.43 [2.32, 5.35] | 8.35 [6.75, 10.74] | 0.58 [0.43, 0.90] | 4.66 [4.29, 5.07] | 4.55 [3.97, 5.35] | 15.11 [11.67, 20.4] |
|  | 40-50 years<br>n = 4442 | 4.31 [2.84, 6.46] | 8.74 [6.82, 11.16] | 0.99 [0.61, 1.75] | 4.77 [4.34, 5.27] | 5.72 [4.78, 7.19] | 17.52 [13.15, 23.26] |
|  | 60-70 years<br>n = 6632 | 4.71 [3.32, 6.53] | 7.92 [6.05, 10.27] | 1.62 [1.03, 2.53] | 4.47 [4.05, 4.94] | 7.52 [6.47, 9.37] | 17.62 [13.54, 23.02] |
| Male | 20-30 years<br>n = 1880 | 2.87 [1.83, 4.53] | 4.74 [3.36, 6.46] | 1.03 [0.69, 1.82] | 6.21 [5.69, 6.80] | 5.36 [4.64, 6.52] | 10.30 [7.11, 14.74] |
|  | 40-50 years<br>n = 5253 | 4.03 [2.89, 5.68] | 4.88 [3.75, 6.38] | 2.71 [1.62, 3.99] | 6.48 [5.92, 7.12] | 7.38 [6.14, 10.41] | 12.76 [9.78, 16.66] |
|  | 60-70 years<br>n = 6702 | 3.77 [2.72, 5.09] | 4.31 [3.28, 5.70] | 3.46 [2.21, 4.78] | 6.21 [5.61, 6.85] | 8.69 [7.38, 11.82] | 12.04 [9.22, 15.44] |
| **C. Age and sex effects** |  |  |  |  |  |  |  |
| **Metric** | **Age Effect Female** | **Age Effect Male** | **Sex Effect Young Adult F/M** | **Sex Effect Older Adult F/M Ratio** |  |  |  |
| ASAT | 1.37 | 1.31 | 1.20 | 1.25 |  |  |  |
| GFAT | 0.95 | 0.91 | 1.76 | 1.84 |  |  |  |

|  |  |  |  |  |
| --- | --- | --- | --- | --- |
| VAT | 2.79 | 3.36 | 0.56 | 0.47 |
| Muscle | 0.96 | 1.00 | 0.75 | 0.72 |
| LFF | 1.65 | 1.62 | 0.85 | 0.87 |
| Total SAT | 1.17 | 1.17 | 1.47 | 1.46 |

*Note: Values in Panel A and B represent median volumes and interquartile ranges (IQR). In Panel C, age effects represent relative changes from age 20-30 to age 60-70, and sex effects represent female-to-male ratios in the 20-30 and 60-70 age group respectively. ASAT = abdominal subcutaneous adipose tissue; GFAT = gluteofemoral adipose tissue; VAT = visceral adipose tissue; LFF = liver fat fraction; Total SAT = total subcutaneous adipose tissue.*

Men had significantly higher VAT and trunk muscle volumes than women in both cohorts (p□<□0.0001 for all comparisons). In NAKO, VAT was 2.69□L (IQR: 1.43– 4.14) in men versus 1.06□L (IQR: 0.59–2.00) in women; in UKB, 3.10□L (IQR: 2.03–4.38) versus 1.47□L (IQR: 0.98–2.28). Trunk muscle volumes showed a similar pattern (NAKO: 6.38□L [IQR: 5.82–7.00] in men vs. 4.74□L [IQR: 4.34–5.21] in women; UKB: 6.21□L [IQR: 5.57–6.91] vs. 4.40□L [IQR: 3.97–4.87]). Conversely, GFAT was substantially higher in women: 8.69□L (IQR: 6.81–11.12) in NAKO and 7.75□L (IQR: 5.93–10.06) in UKB, compared to 4.82□L (IQR: 3.69–6.26) and 4.16□L (IQR: 3.13–5.55) in men, respectively (p□<□0.0001 for all comparisons).

These sex-based differences were evident across all BMI categories (Figure 2c) and persisted among individuals with normal weight (Table 3A), suggesting that compartmental fat and muscle distribution varies systematically by sex and age, independent of BMI.

### Correlations among adjusted body composition parameters and associations with lifestyle factors

We derived residualised body composition parameters adjusted for age, sex (for combined analyses), height, and BMI using linear regression models to examine relationships while holding these covariates constant. The resulting correlation matrix of adjusted values is shown in Figure 2b and Supplementary Table S3. ASAT-adj and GFAT-adj were positively correlated (r = 0.56 in men), whereas muscle-adj showed inverse correlations with both subcutaneous fat compartments across sexes. VAT-adj and LFF-adj were positively correlated in both men (r = 0.35) and women (r = 0.43), indicating shared variance not explained by anthropometric covariates.

In NAKO participants, physical activity was associated with body composition patterns characterized by lower VAT and higher trunk muscle volume (Supplementary Table S4). Compared to participants classified as insufficiently active (<600 MET min/week), those in the moderate activity group (1000–2999 MET min/week) had significantly lower VAT Z-scores (–0.130, 95% CI [–0.152, –0.107], p < 0.0001). Sufficient activity levels (600–999 MET min/week) were associated with smaller reductions (–0.037, 95% CI [–0.069, –0.005], p = 0.024), while participants in the highest category (≥3000 MET min/week) showed only a slightly greater reduction (–0.156, 95% CI [–0.176, –0.136], p < 0.0001), with diminishing additional effect at the upper end of the activity range. Hand grip strength was also independently associated with lower VAT and higher trunk muscle volume.

Among UKB participants, current smoking was associated with lower adjusted ASAT (–0.052, 95% CI [–0.083, –0.020], p = 0.0019) and GFAT (–0.072, 95% CI [–0.107, – 0.036], p = 0.0001), and higher VAT (0.046, 95% CI [0.004, 0.089], p = 0.0426) and LFF (0.085, 95% CI [0.022, 0.148], p = 0.018). These findings reflect altered fat distribution among current smokers; however, causal inference is not possible from these cross-sectional associations (Supplementary Fig. S3). Associations between alcohol intake frequency and body composition parameters are reported in Figure 2, Supplementary Table S5 and Supplementary Figure S3d as well as Supplementary Results.

### Associations between body composition phenotypes and cardiometabolic conditions

Associations between body composition parameters and clinical conditions were estimated per 1 SD higher compartment volume, adjusted for age, sex, height, and BMI, reflecting variation in fat and muscle distribution among individuals with comparable body size and demographics. Higher GFAT-adj was consistently associated with lower prevalence of type 2 diabetes (OR = 0.69, 95% CI 0.66–0.71), coronary artery disease (OR = 0.88, 95% CI 0.81–0.95), hypertension (OR = 0.88, 95% CI 0.86–0.90), and hyperlipidemia (OR = 0.87, 95% CI 0.85–0.89) (Figure 3b). In contrast, ASAT-adj showed modestly higher odds of hypertension (OR = 1.05, 95% CI 1.03–1.08), type 2 diabetes (OR = 1.06, 95% CI 1.02–1.11), and hyperlipidemia (OR = 1.11, 95% CI 1.08–1.13) (Figure 3a). VAT-adj was most strongly associated with prevalence of cardiometabolic conditions: OR = 1.12 (95% CI 1.05–1.19) for coronary artery disease, OR = 1.20 (95% CI 1.08–1.33) for heart failure, OR = 1.19 (95% CI 1.16–1.22) for hypertension, OR = 1.19 (95% CI 1.14–1.24) for type 2 diabetes, and OR = 1.29 (95% CI 1.26–1.32) for hyperlipidemia (Figure 3c). As these models adjust for BMI and height, increases in a given compartment reflect substitution for other compartments at constant body size, rather than effects of absolute volume.^24,25^ Observed associations for higher GFAT-adj reflect variation in gluteofemoral fat relative to other compartments, rather than direct evidence for beneficial properties of GFAT itself. These cross-sectional associations remained significant in models including interaction terms with BMI (Figure 3g) and were also evident in normal-weight individuals.

**Figure 3.**
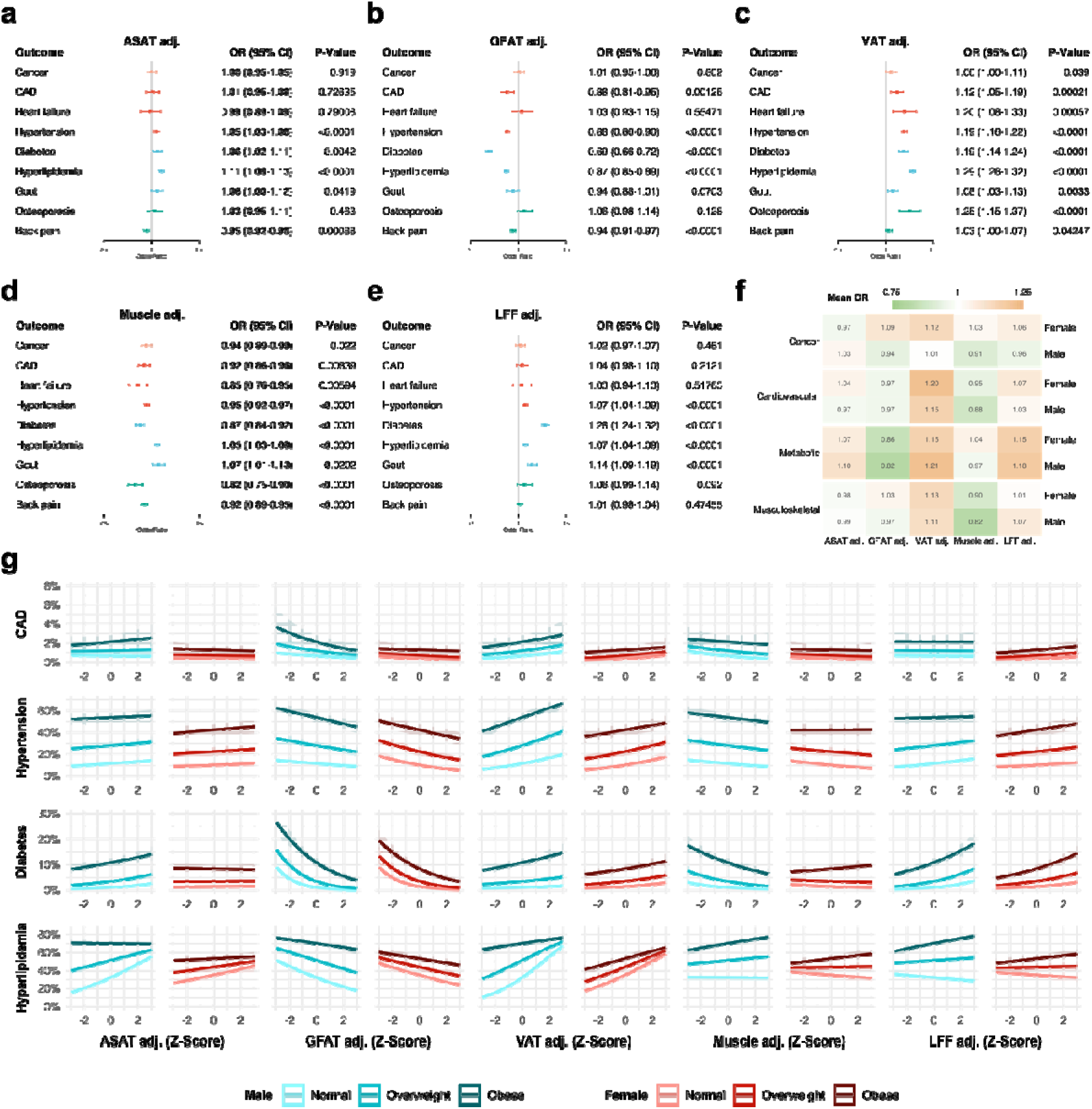
Associations between body composition parameters and clinical conditions. (a–e) Associations between adjusted body composition parameters (BCPs) and the prevalence of clinical conditions, presented as odds ratios (OR) derived from generalized mixed-effects logistic regression models. P-values for fixed effects were derived using Wald z-statistics. Models included adjusted BCPs, age, BMI, and sex, with imaging centre included as a random effect. Cancer data were available only in UKB participants. Colours indicate clinical condition categories. (f) Heatmap of average odds ratios for each clinical condition group, based on sex-stratified models (detailed results in Supplementary Figure S5). (g) Estimated cross-sectional prevalence of selected conditions across adjusted BCP Z-scores, shown for BMI reference values of 20 kg/m² (normal), 27.5 kg/m² (overweight), and 35 kg/m² (obese). Estimates are based on marginalized means from sex-stratified generalized mixed-effects logistic regression models including interaction terms between BMI and each BCP, with imaging centre as a random effect. These models describe prevalence variation and should not be interpreted as predictive of future risk.

LFF-adj was positively associated with hypertension (OR = 1.07, 95% CI 1.04–1.09), type 2 diabetes (OR = 1.28, 95% CI 1.24–1.32), and hyperlipidemia (OR = 1.07, 95% CI 1.04–1.09) (Figure 3e). In BMI-stratified models, an inverse association with hyperlipidemia was observed in normal-weight individuals, indicating potential variation by BMI category in these associations (Figure 3g). Muscle-adj showed inverse associations with several diseases/clinical conditions: type 2 diabetes (OR = 0.87, 95% CI 0.84–0.92), heart failure (OR = 0.85, 95% CI 0.76–0.95), and hypertension (OR = 0.95, 95% CI 0.92–0.97) (Figure 3d).

Non-cardiometabolic associations followed similar trends. Gout was associated with higher ASAT-adj, VAT-adj, muscle-adj, and LFF-adj, and lower GFAT-adj. Osteoporosis showed opposing associations with VAT-adj (OR = 1.25, 95% CI 1.15– 1.37) and muscle-adj (OR = 0.82, 95% CI 0.75–0.90). Back pain was modestly less common with higher ASAT-adj (OR = 0.95, 95% CI 0.92–0.98), GFAT-adj (OR = 0.94, 95% CI 0.91–0.97), and muscle-adj (OR = 0.92, 95% CI 0.89–0.95).

Distinct disease-specific body composition phenotypes were observed. Individuals with type 2 diabetes had elevated VAT-adj (+0.45□±□1.31), reduced GFAT-adj (–0.45□±□1.18), and elevated LFF-adj (+0.60□±□1.61). Hypertensive individuals had higher VAT-adj (+0.20□±□1.19) and slightly lower GFAT-adj (–0.11□±□1.07), compared to healthy participants (VAT-adj: –0.16□±□0.86; GFAT-adj: +0.07□±□1.00) (Table 4). Based on these patterns, we defined an “unfavorable” body composition profile as +1 SD in ASAT-adj, VAT-adj, and LFF-adj, and –1 SD in GFAT-adj and muscle-adj. Predicted clinical conditions across age are shown for mean and unfavorable phenotypes in Figure 4d–f.

**Table 4.** Body composition Z-scores by clinical condition. This table presents mean Z-scores of adjusted body composition parameters (abdominal subcutaneous adipose tissue [ASAT], gluteofemoral adipose tissue [GFAT], visceral adipose tissue [VAT], trunk muscle volume, and liver fat fraction [LFF]) across multiple clinical conditions, as determined in the provided dataset. Each row corresponds to a specific disease/condition category, listing the number of individuals (n), followed by the mean Z-score and standard deviation (with standard error in parentheses) for each parameter. Positive Z-scores indicate higher relative values compared to the average total population, whereas negative values indicate lower relative values.

| Clinical Condition | n | Mean Z-Scores |  |  |  |  |
| --- | --- | --- | --- | --- | --- | --- |
|  |  | ASAT adj. | GFAT adj. | VAT adj. | Muscle adj. | LFF adj. |
| Cardiovascular |  |  |  |  |  |  |
| CAD | 917 | -0.09 ± 1.06 (0.03) | -0.07 ± 0.96 (0.03) | 0.23 ± 1.29 (0.04) | 0.01 ± 1.09 (0.04) | 0.06 ± 1.25 (0.04) |
| Heart failure | 365 | 0.04 ± 1.09 (0.06) | 0.00 ± 1.22 (0.06) | 0.20 ± 1.13 (0.06) | -0.12 ± 1.01 (0.05) | 0.12 ± 1.16 (0.06) |
| Hypertension | 10,869 | 0.02 ± 1.11 (0.01) | -0.11 ± 1.07 (0.01) | 0.20 ± 1.19 (0.01) | 0.02 ± 1.09 (0.01) | 0.13 ± 1.29 (0.01) |
| Metabolic |  |  |  |  |  |  |
| Diabetes | 2,290 | -0.05 ± 1.25 (0.03) | -0.45 ± 1.18 (0.02) | 0.45 ± 1.31 (0.03) | 0.05 ± 1.17 (0.02) | 0.60 ± 1.61 (0.03) |
| Hyperlipidaemia | 20,570 | 0.04 ± 1.02 (0.01) | -0.10 ± 1.00 (0.01) | 0.16 ± 1.05 (0.01) | 0.07 ± 1.02 (0.01) | 0.10 ± 1.14 (0.01) |
| Gout | 1,268 | -0.04 ± 1.21 (0.03) | -0.19 ± 1.04 (0.03) | 0.41 ± 1.30 (0.04) | 0.14 ± 1.17 (0.03) | 0.36 ± 1.44 (0.04) |
| Other |  |  |  |  |  |  |
| Back pain | 6,070 | -0.03 ± 1.03 (0.01) | -0.05 ± 1.04 (0.01) | 0.03 ± 1.02 (0.01) | -0.02 ± 0.98 (0.01) | 0.03 ± 1.07 (0.01) |
| Cancer | 2,147 | 0.02 ± 0.97 (0.02) | 0.03 ± 1.02 (0.02) | -0.01 ± 1.01 (0.02) | -0.08 ± 0.96 (0.02) | 0.01 ± 1.03 (0.02) |
| Osteoporosis | 875 | 0.11 ± 0.87 (0.03) | 0.04 ± 1.01 (0.03) | 0.09 ± 0.91 (0.03) | -0.05 ± 0.82 (0.03) | 0.10 ± 1.04 (0.04) |
| Reference |  |  |  |  |  |  |
| Healthy | 6,393 | -0.01 ± 0.96 (0.01) | 0.07 ± 1.00 (0.01) | -0.16 ± 0.86 (0.01) | -0.08 ± 0.96 (0.01) | -0.05 ± 0.82 (0.01) |

*Note: Values represent mean ± standard deviation with standard error in parentheses. Positive values indicate higher relative values compared to the total population. ASAT = abdominal subcutaneous adipose tissue; GFAT = gluteofemoral adipose tissue; VAT = visceral adipose tissue; LFF = liver fat fraction; adj. = adjusted.*

**Figure 4.**
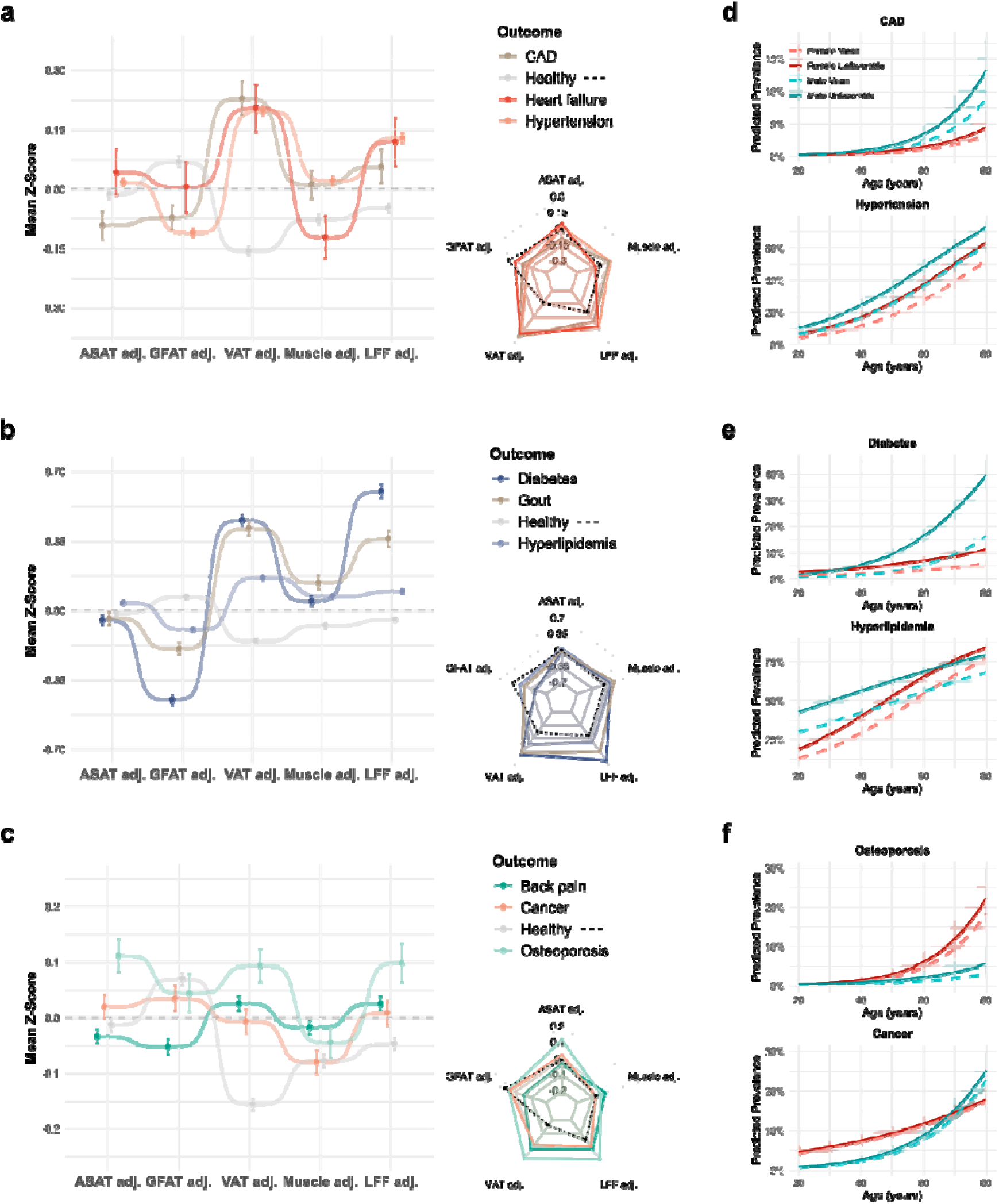
Body composition phenotypes and their associations with prevalence of cardiometabolic, cancer, and related clinical conditions. (a–c) Mean Z-scores of adjusted body composition parameters (BCPs) for participants with different clinical conditions, displayed using dot-whisker and spider plots. Values reflect mean (± standard error) adjusted BCP Z-scores for each condition. "Healthy" was defined as participants without any of the assessed conditions. (d–f) Estimated cross-sectional prevalence of cardiometabolic, cancer, and related clinical conditions across age for two standardized body composition phenotypes: “unfavourable” (defined as +1 SD in ASAT, VAT, and LFF; –1 SD in GFAT and trunk muscle volume) and “mean” (Z = 0 for all BCPs). Estimates are based on marginalized predictions from sex-stratified generalized linear mixed-effects models adjusted for age, BMI, and adjusted BCPs, with imaging centre modelled as a random effect. These models describe variation in prevalence, not risk or prognosis. Cancer data were available only in UKB participants.

### Associations between body composition phenotypes and cancer prevalence

The relationship between body composition and cancer was evaluated in UKB participants with available diagnosis data. Associations were estimated per 1 SD higher compartment volume, adjusted for age, sex, height, and BMI, reflecting differences in fat and muscle distribution among individuals with similar body size. Higher VAT-adj was associated with slightly higher cancer prevalence (OR = 1.06, 95% CI 1.00–1.11, p = 0.036; Figure 3c). This association was evident in women (OR = 1.12, 95% CI 1.05–1.21, p = 0.0012), with no significant association in men (Supplementary Figure S5). GFAT-adj was not related to cancer in the full sample (OR = 1.01, 95% CI 0.95–1.06, p = 0.80), but showed a positive association in women (OR = 1.09, 95% CI 1.01–1.17, p = 0.027). Muscle-adj was inversely associated with cancer prevalence overall (OR = 0.94, 95% CI 0.89–0.99, p = 0.022), with this association largely driven by men (OR = 0.91, 95% CI 0.85–0.99, p = 0.026). LFF-adj was not significantly associated with cancer (OR = 1.02, 95% CI 0.97–1.07, p = 0.46).

Participants with cancer exhibited modest differences in adjusted body composition parameters compared to healthy controls (Table 4), including slightly higher VAT-adj (–0.01□±□1.01 vs. –0.16□±□0.86) and ASAT-adj (+0.02□±□0.97 vs. – 0.01□±□0.96). GFAT-adj was similar between groups (+0.03□±□1.02 in cancer vs. +0.07□±□1.00 in controls), and muscle-adj was identical (–0.08□±□0.96 in both groups). Sex-stratified analyses further revealed that, at a given BMI, women with cancer had higher visceral adipose tissue than controls, while men with cancer had lower trunk muscle volume, suggesting sex-specific shifts in body composition (Supplementary Table S6).

Age-stratified models showed pronounced sex differences in predicted cancer prevalence (Table 5). In men, prevalence increased from 1.49% (95% CI 0.49–3.63) at ages 20–40 to 15.42% (95% CI 7.65–30.05) at ages 61–80. In women, corresponding estimates were 5.90% (95% CI 3.54–9.03) and 14.58% (95% CI 10.46–20.81), resulting in a female-to-male ratio of 3.95 in early adulthood that declined with age. This pattern coincided with age-related increases in VAT, particularly among men (Table 3), although causality cannot be inferred from these cross-sectional data.

**Table 5.** Predicted prevalence of cardiometabolic, cancer, and related clinical conditions by sex and age. This table presents the predicted prevalence of cardiometabolic, cancer, and related clinical conditions for three age groups (20–40, 41–60, 61–80 years) in females and males.

| Clinical Condition | Sex | Age Group | Predicted Prevalence (%) | Sex Ratio (F/M) |
| --- | --- | --- | --- | --- |
| Cardiovascular Conditions |  |  |  |  |
| CAD | Female | 20-40 years | 0.23 (0.06–0.65) | 0.61 |
|  |  | 41-60 years | 0.75 (0.25–1.99) | 0.47 |
|  |  | 61-80 years | 2.40 (0.86–6.90) | 0.38 |
|  | Male | 20-40 years | 0.37 (0.09–1.19) | 1.00 |
|  |  | 41-60 years | 1.59 (0.43–4.73) | 1.00 |
|  |  | 61-80 years | 6.33 (1.87–18.17) | 1.00 |
| Hypertension | Female | 20-40 years | 9.55 (3.62–20.77) | 0.68 |
|  |  | 41-60 years | 23.29 (10.34–42.89) | 0.74 |
|  |  | 61-80 years | 45.75 (24.85–68.59) | 0.82 |
|  | Male | 20-40 years | 14.03 (5.71–28.28) | 1.00 |
|  |  | 41-60 years | 31.58 (15.52–52.83) | 1.00 |
|  |  | 61-80 years | 55.83 (34.23–76.33) | 1.00 |
| Metabolic Conditions |  |  |  |  |
| Diabetes | Female | 20-40 years | 2.66 (1.10–5.40) | 1.24 |
|  |  | 41-60 years | 4.35 (1.97–8.58) | 0.63 |
|  |  | 61-80 years | 6.96 (3.20–13.99) | 0.37 |
|  | Male | 20-40 years | 2.15 (0.40–6.65) | 1.00 |
|  |  | 41-60 years | 6.87 (1.53–19.04) | 1.00 |
|  |  | 61-80 years | 18.92 (5.08–45.10) | 1.00 |
| Hyperlipidaemia | Female | 20-40 years | 24.65 (11.13–44.21) | 0.58 |
|  |  | 41-60 years | 47.90 (27.47–69.26) | 0.85 |
|  |  | 61-80 years | 71.60 (51.72–86.63) | 1.04 |
|  | Male | 20-40 years | 42.85 (27.61–59.48) | 1.00 |
|  |  | 41-60 years | 56.32 (40.53–71.70) | 1.00 |
|  |  | 61-80 years | 68.65 (53.88–81.54) | 1.00 |
| Other Conditions |  |  |  |  |
| Osteoporosis | Female | 20-40 years | 0.67 (0.17–2.09) | 1.68 |
|  |  | 41-60 years | 2.99 (0.86–8.53) | 2.74 |
|  |  | 61-80 years | 11.90 (3.79–30.04) | 4.13 |
|  | Male | 20-40 years | 0.40 (0.09–1.44) | 1.00 |
|  |  | 41-60 years | 1.09 (0.29–3.72) | 1.00 |
|  |  | 61-80 years | 2.88 (0.79–10.03) | 1.00 |
| Cancer | Female | 20-40 years | 5.90 (3.54–9.03) | 3.95 |
|  |  | 41-60 years | 9.43 (6.37–13.43) | 1.86 |
|  |  | 61-80 years | 14.58 (10.46–20.81) | 0.95 |
|  | Male | 20-40 years | 1.49 (0.49–3.63) | 1.00 |
|  |  | 41-60 years | 5.08 (2.06–10.81) | 1.00 |
|  |  | 61-80 years | 15.42 (7.65–30.05) | 1.00 |

*Note: Mean predicted prevalence was multiplied by 100 and is presented with the smallest lower confidence bound and largest upper confidence bound observed in each group. Sex Ratio (F/M) is the female mean divided by the male mean prevalence for the matching age range.*

## Discussion

Adipose tissue distribution, rather than BMI, is associated with cardiometabolic and cancer prevalence, as revealed by our comprehensive MRI analysis of 45,851 participants across two European cohorts. Elevated VAT was consistently associated with prevalence of cardiometabolic conditions even in normal-weight individuals, with significantly higher odds of conditions like coronary artery disease, hypertension, and type 2 diabetes.

By contrast, GFAT was inversely associated with multiple conditions. However, given that associations were estimated at a constant body size, these findings may also reflect a redistribution of fat from visceral compartments rather than a depot-specific protective effect of gluteofemoral fat. Utilizing a fully automated deep-learning pipeline, we generated volumetric measurements in approximately one minute per scan, demonstrating the feasibility of scalable and reproducible body composition biomarkers for population-level research and risk stratification. Characteristic body composition phenotypes were identified for specific diseases; for example, patients with diabetes exhibited elevated VAT, reduced GFAT, and increased liver fat, adjusted for BMI.

The distinct associations of VAT and GFAT with prevalence of clinical conditions observed in our analysis support previously described biological differences between these adipose depots in smaller clinical and experimental studies. VAT exerts adverse cardiometabolic effects through well-documented mechanisms involving insulin resistance, dyslipidaemia, and chronic systemic inflammation.^26–34^ Our results further indicate that, among adults of similar BMI, those with relatively higher GFAT and lower VAT have a lower prevalence of cardiometabolic conditions. While these observations align with mechanistic hypotheses regarding metabolic buffering and depot-specific adipokine secretion,^35^ our study underscores the importance of fat distribution for individuals of similar body size, rather than effects of absolute changes in single depots.

Similarly, trunk muscle volume showed inverse associations with the prevalence of cardiometabolic conditions after adjustment for age and BMI, with lower odds of type 2 diabetes, among others, likely reflecting variation in muscle mass at a given body size. This inverse association hints at muscle tissue’s critical functions in glucose homeostasis, insulin sensitivity, and inflammatory modulation.^35^ Associations were more pronounced in men, whereas in women, associations were primarily observed for musculoskeletal conditions such as osteoporosis and back pain. These findings underscore the potential clinical relevance of preserved muscle mass through improved glucose disposal, anti-inflammatory effects, and mechanical loading.^36,37^ Cancer-related findings from UKB add to existing evidence that body composition is associated with differences in cancer prevalence. With MRI phenotyping, we identified sex-specific associations not captured by BMI. Higher VAT-adj was associated with increased cancer prevalence in women, with no such association in men. This female-specific pattern may reflect differences in hormonal, adipokine, or inflammatory profiles.^38^ While GFAT-adj was inversely associated with prevalence of cardiometabolic conditions, it showed a positive association with cancer prevalence in women, suggesting that adipose tissue distribution may reflect sex-specific variation in cancer susceptibility patterns. Muscle-adj was inversely associated with cancer prevalence in men, while no association was observed in women, suggesting sex-specific metabolic or inflammatory contributions.

Disease-specific body composition phenotypes provided additional nuance to our understanding of pathophysiological relationships. Cardiometabolic conditions exhibited pronounced deviations in body composition parameters, while cancer patients showed more subtle alterations that varied by sex. These MRI-derived phenotypes may improve clinical stratification beyond traditional anthropometric measures.^39,40^ Age- and sex-stratified models also revealed clinically relevant patterns. For example, women with high VAT and low muscle mass reached hyperlipidaemia prevalence nearly a decade earlier than men with average body composition phenotypes. Such observations support the added value of regional body composition phenotyping for clinical risk assessment and highlight the heterogeneity of disease burden within BMI categories.

Our analysis of age-related changes in body composition revealed differential trajectories by sex and tissue type. VAT accumulation increased with age, particularly in men, while GFAT was relatively preserved or declined modestly. These shifts paralleled age-related patterns of prevalence of clinical conditions and underscore the importance of longitudinal assessment of body composition. Physical activity showed a non-linear association with VAT and muscle mass, with the greatest benefits observed at moderate activity levels and diminishing additional effects at high activity levels. These findings may inform targeted interventions to mitigate age-related shifts in body composition associated with prevalence of clinical conditions.

Altogether, these findings emphasize critical limitations of BMI as an obesity metric, as it fails to differentiate metabolically detrimental visceral fat from protective subcutaneous fat compartments or beneficial muscle mass. Our results demonstrate that, among individuals with comparable BMI, substantial heterogeneity in disease prevalence is explained by differences in regional fat and muscle distribution. Normal-weight individuals with high VAT showed comparable prevalence of clinical conditions to those with obesity, while those with obesity but lower VAT and preserved muscle mass showed lower observed prevalence of clinical conditions.^41,42^ MRI-derived body composition phenotypes were cross-sectionally associated with cardiometabolic and cancer prevalence, independent of traditional anthropometric and behavioural covariates, enabling more granular characterization of adiposity-related disease patterns. The observed heterogeneity within BMI categories highlights the potential value of MRI-based phenotyping for refining current obesity classification systems and for characterizing prevalence of clinical conditions in epidemiologic research.

This study has limitations. The observational, cross-sectional design inherently limits any inference regarding directionality or causality. Reverse causation is possible, particularly for conditions such as cancer or heart failure, where disease may influence body composition. Selection biases due to voluntary participation may influence representativeness, with both cohorts comprising predominantly middle-aged individuals of European descent from urban environments. Despite rigorous harmonization efforts, residual methodological differences between cohorts could affect comparability. While MRI provides accurate body composition quantification, clinical implementation depends on the widespread availability and accessibility of imaging resources. Future research should prospectively validate these findings, evaluate cost-effectiveness, and explore longitudinal body composition changes alongside functional metrics. Since all associations observed in this study were adjusted for BMI and height, they reflect variation in fat and muscle distribution at a constant body size and should be interpreted as reflecting differences in fat and muscle distribution, rather than the effect of absolute compartment size.

Our findings demonstrate the utility of MRI-based body composition phenotyping for epidemiological surveillance and stratification of prevalence of clinical conditions. Regional adiposity and muscle phenotypes were distinctly associated with cardiometabolic and cancer conditions across BMI strata, offering greater granularity than conventional anthropometric measures. While causal inference is limited by the cross-sectional design, these results provide a basis for hypothesis generation and support prospective studies to assess whether modifying specific body composition compartments improves clinical conditions in interventional or longitudinal settings.

## Methods

This study was conducted under data access applications NAKO-836 and UKB-34479 for the German National Cohort and UK Biobank, respectively. The full protocols for NAKO and UKB are available online (NAKO: https://nako.de/wp-content/uploads/NAKO-Wissenschaftliches-Konzept.pdf; UKB: https://www.ukbiobank.ac.uk/media/gnkeyh2q/study-rationale.pdf). Informed consent was obtained from all participants from NAKO and UKB. In addition, we received local IRB approval (2024-479-S-SB).

### Study cohorts

Data for this study were derived from the NAKO and UKB cohorts, with participant selection outlined in Supplementary Figure S8.

The NAKO is a population-based, multi-centric, prospective study of 205,217 individuals aged 19–74 years. Data collection for the baseline examination was performed between 2014 and 2019 at 18 study centers.^21,43^ Whole-body 3T MRI scans were acquired for 30,861 participants at five dedicated centres across Germany using identical Magnetom Skyra scanners (Siemens Healthineers, Erlangen, Germany). MRI and outcome data were available for 30,397 participants. For body composition assessment, in-phase gradient echo (GRE) images were processed by stitching together four separate image sections using an in-house tool.

Participants were excluded if required GRE image types (in-phase, out-of-phase, fat-only, water-only) or expected partition numbers (4 for NAKO) were missing, or if image dimensions were inconsistent. Next, 137 participants with liver fat fraction (LFF) > 0.6 over larger regions, indicative of fat–water swap artifacts, were excluded. Following additional exclusions due to missing anthropometric (n = 1), disease status (n = 787), or health behaviour data (n = 2,237), 26,877 individuals (14,969 males and 11,908 females) were included in the final analysis.

The UKB is a large-scale biomedical database consisting of over 500,000 participants aged 40–69 years, with baseline data collected between 2006 and 2010. For this study, MRI imaging and outcome data from 19,512 participants scanned at two imaging centres were available. Dixon images (in-phase GRE) were processed by stitching together six separate image sections using an in-house tool. We excluded 91 participants for incorrect image structure or dimension, and 257 with LFF > 0.6 due to fat–water swap artifacts. A further 190 were excluded for missing health behaviour data, resulting in 18,974 participants (9,094 males and 9,880 females) in the final dataset.

### Classification of health behaviours and clinical conditions

Classification was implemented to standardize the assessment of health behaviour indicators, covariates, and clinical condition outcomes across datasets. In the NAKO cohort, hand grip strength was measured as the maximum isometric grip strength value of at least two measurement values using a JamarPlus+ hand dynamometer (Sammons Preston, Rolyon, Bolingbrook, IL, USA), while physical activity was quantified using self-reported weekly metabolic equivalent of task (MET) minutes. Based on the minimum recommendations from the World Health Organization (WHO, 2021) and evidence indicating benefits beyond, physical activity was categorized into four levels: insufficient (<600 MET minutes/week), sufficient (600–999 MET minutes/week), moderate (1000–2999 MET minutes/week), and high (≥3000 MET minutes/week). For UKB participants, data on lifestyle factors, including smoking status and alcohol intake frequency, were available and incorporated to analyze their influence on body composition.

Clinical condition classification focused on key metabolic and cardiovascular diseases (coronary artery disease, type 2 diabetes, hypertension), cancer, as well as related risk factors and clinical traits such as hyperlipidemia, osteoporosis, and back pain. For NAKO participants, coronary artery disease (CAD) was defined based on a self-reported history of heart attack, known constriction of the coronary arteries, or angina pectoris. Type 2 diabetes was identified either through self-report or, for NAKO participants, by an HbA1c level of ≥6.5% measured within three months prior to imaging. Hyperlipidemia was classified using self-reported data on high cholesterol (UKB), and high cholesterol or triglycerides (NAKO). Additional laboratory thresholds were applied for NAKO participants not currently on lipid-lowering medication: total cholesterol ≥240 mg/dl, LDL cholesterol ≥160 mg/dl, or triglycerides ≥200 mg/dl. Other clinical conditions, such as hypertension, osteoporosis, and back pain, were classified based on self-reported medical history collected during participant assessment. Data regarding history of cancer was only available in UKB participants. Cancer status in UKB was determined from self-report and linked health records (ICD-10 codes C00–C97). For clarity, we use the term ‘clinical condition’ throughout this manuscript to encompass both diagnosed diseases (e.g., coronary artery disease, type 2 diabetes, cancer) and related risk factors or clinical traits (e.g., hyperlipidemia, back pain, osteoporosis) as defined in the study cohorts.

### Deep learning-based segmentation of body composition parameters

Body composition parameters were extracted using *MRSegmentator*^44^, which was extended to include segmentations for SAT, ASAT, GFAT, VAT, and trunk musculature. ASAT was delineated from the top of the T9 vertebra to the top of the gluteus maximus dorsally and the pubic symphysis ventrally, with the posteroanterior boundary defined by the iliac bone. GFAT was segmented continuously from the ASAT down to the knee joint line to capture lower body fat distribution.

Segmentation used an nn-U-Net with a learning rate of 3 × 10⁻□, 1,000 epochs, a patch size of 192 × 192 × 64 voxels and on-the-fly affine, elastic and gamma-intensity augmentations. It was validated using 5-fold cross-validation (VAT Dice 0.83 ± 0.09, GFAT Dice 0.95 ± 0.03).

We employed a human-in-the-loop training strategy. Initially, 20 manually segmented MRIs per cohort (NAKO and UKB; 40 total) were used to train a preliminary nnU-Net model.^45^ These scans were selected to reflect variation in body habitus and image quality. The model was then applied to additional scans, with pre-segmentations manually corrected and used for retraining. Additionally, UKB in-phase annotations were registered to the corresponding opposed-phase, fat-only, and water-only images to enable full multi-sequence generalizability. In total, 250 curated segmentations (50 NAKO, 200 UKB) were used for final model development. This iterative approach enabled efficient expansion of the training set while improving generalizability.

Voxel volumes of segmented tissues were extracted for further analysis, and liver fat fractions (LFF) were calculated from fat-only and water-only signal intensities.^46,47^ Fat–water swap artifacts were not corrected algorithmically. Instead, scans with LFF ≥ 0.6 over larger liver regions were excluded following visual confirmation. When smaller regions exceeded this threshold, these were masked to prevent bias in LFF estimation.

Systematic spot checks were performed to assess segmentation quality, focusing on anatomical plausibility and outlier volumes. A total of 76 participants (<0.2%) were excluded due to segmentation anomalies, including truncated anatomy or misregistration. These errors were randomly distributed across scanners and cohorts, with no evidence of systematic bias.

### Data processing and statistical analysis

Data were harmonised with ComBat,^48,49^ modelling scanner vendor, field strength and echo time as batch variables while preserving age, sex and BMI as biological covariates (Supplementary Figure S9). Participants with missing covariate or outcome data were excluded listwise from each model. No imputation was performed. Individual muscle compartments were separately assessed as illustrated in Supplementary Figure S10. Participants with missing covariate or outcome data were excluded from the respective models; no imputation was applied.

Linear mixed-effects models were used to examine associations between lifestyle factors and MRI-derived body composition parameters. In these models, fixed effects included z-standardized age, height, and BMI, sex (in combined analyses), along with z-standardized maximum hand grip strength and physical activity levels for the NAKO cohort or smoking status and alcohol intake frequency for the UKB cohort. The imaging centre was incorporated as a random effect to account for site-specific variability.

To investigate body composition independent of age, height, BMI, and sex (in combined analyses), adjusted body composition parameters were derived from the residuals of respective linear regression models. To assess associations between body composition and status of clinical conditions, generalized linear mixed-effects models were employed. These models included the adjusted body composition parameters, age, BMI, and sex (in combined analyses) as fixed effects, with the imaging centre again modelled as a random effect. From these models, odds ratios and predicted prevalences were calculated to describe cross-sectional associations with clinical conditions.

P-values for fixed effects of all models were derived using Wald z-statistics, calculated as the ratio of each coefficient estimate to its standard error, assuming asymptotic normality.

Because BMI and height were included as covariates in all models, effect estimates for body composition compartments should be interpreted as reflecting differences in regional fat and muscle distribution for individuals with similar body size, rather than as effects of absolute changes in a single compartment. Increases in one compartment are therefore associated with a redistribution from other compartments at constant BMI and height.

Statistical analyses were performed in *R Version 4.4.3* using the package *lme4*^50^ for modelling, *ggcorrplot* for generating correlation matrices, *forestplot* and *pheatmap* for visualizing odds ratios, *ggeffects*^51^ for computing and plotting marginal predictions, and *ggbump* and *fmsb* for constructing body composition profile plots. Plots of cohort characteristics and harmonized body composition metrics were created using the Python package seaborn.

## Data Availability

The data analyzed in this study were obtained from the German National Cohort (NAKO Gesundheitsstudie) and UK Biobank. Access to these datasets is subject to the respective data access policies and requires a formal application to the data custodians. Further details regarding data access procedures can be found on the NAKO (https://nako.de/) and UK Biobank (https://www.ukbiobank.ac.uk/) websites. The authors do not have permission to share the raw data directly.

## Supplementary Results

We assessed associations between self-reported alcohol intake frequency and adjusted body composition parameters in 18,974 UKB participants using linear mixed-effects models adjusted for age, height, BMI, and sex (in combined analyses), with imaging center as a random effect. Results are presented in Supplementary Table S5.

Alcohol intake showed inverse associations with subcutaneous fat compartments. Compared to abstainers, participants consuming alcohol 3–4 times per week had lower adjusted ASAT (β = –0.054, 95% CI [–0.080, –0.027], p = 0.0001) and GFAT (β = –0.060, 95% CI [–0.090, –0.030], p = 0.0001). A similar inverse association was observed for daily alcohol intake. In contrast, VAT and liver fat fraction (LFF) were not consistently associated with alcohol frequency.

Notably, alcohol intake frequency was positively associated with adjusted trunk muscle volume, particularly in the 3–4 times per week group (β = 0.093, 95% CI [0.073–0.113], p < 0.0001).

These cross-sectional associations may reflect complex behavioral or health-related confounding. Specifically, individuals who abstain from alcohol may represent a heterogeneous group that includes former drinkers and individuals with underlying health conditions or comorbidities, which could confound associations with muscle mass or adiposity. These findings should be interpreted as descriptive associations only. Alcohol-related results were not adjusted for potential confounders such as diet, education, medication use, or comorbidity burden, limiting interpretability. These exploratory analyses therefore require confirmation in longitudinal cohorts.

## Supplementary Material and Methods

### Deep learning segmentation performance

Whole-body MRI scans were segmented using *MRSegmentator*^44^, a deep learning-based segmentation tool extended in this study to include additional body compartments: subcutaneous adipose tissue (SAT), abdominal subcutaneous adipose tissue (ASAT), gluteofemoral adipose tissue (GFAT), visceral adipose tissue (VAT), and trunk musculature. Figure 1c illustrates segmentation outputs, showing fat depots and musculature including the rectus abdominis (RAM), lateral abdominal (LAM), and quadratus lumborum (QLM) muscles. Liver fat fraction (LFF) was calculated using fat-only and water-only MRI signal intensities (see lower panel, Figure 1c).

Segmentation accuracy was evaluated using 5-fold participant-stratified cross-validation with Dice similarity coefficients (DSC) as the primary metric (Figure 1d). High segmentation accuracy was achieved for subcutaneous compartments (SAT: 0.94, ASAT: 0.95, GFAT: 0.96). GFAT segmentation was limited to UKB due to superior-inferior truncation in NAKO sequences above the hip. VAT segmentation achieved a lower DSC (0.85), reflecting its greater anatomical complexity. Muscle segmentations were consistent across sides and imaging protocols (RAM and QLM: 0.87–0.88; LAM: 0.92–0.93). Importantly, segmentation quality was comparable between T2-HASTE images from NAKO and Dixon-based sequences from UKB (Table S1).

### Segmentation failure analysis

To identify rare failure cases and assess robustness at the extremes, we implemented two complementary quality control procedures: volumetric outlier inspection and ensemble-based uncertainty estimation.

For volumetric inspection, we identified the highest and lowest volume segmentations for VAT, GFAT, SAT, combined musculature, and liver in both cohorts. Visual review confirmed accurate VAT, muscle, and liver segmentations across all extremes. SAT was also correctly segmented in low-BMI participants. However, SAT volume was underestimated in nine participants with extreme obesity (BMI > 40 kg/m²), where body girth exceeded the MRI field of view. GFAT segmentation was consistently accurate in UKB. In contrast, in NAKO, several low-volume scans, primarily in very lean participants, showed under- or over-segmentation near the thighs, likely due to insufficient contrast in the GFAT layer at low fat volumes.

To further probe segmentation reliability, we used a model-discordance heuristic based on the observation by Kofler et al.^48,49^ that inter-model disagreement correlates with segmentation uncertainty. We randomly selected 1,000 participants from each cohort to limit computational and environmental costs, then applied two nnU-Net sub-models trained on different folds. Dice similarity between model outputs was used to identify the lowest-agreement cases for manual review.

Most discrepancies involved subtle boundary shifts between GFAT, SAT, and ASAT, where anatomical transitions are gradual and poorly demarcated. These variations were deemed acceptable on visual inspection. No systematic errors were observed in the UKB cohort. In NAKO, consistent with volumetric findings, the only recurrent issue was under-segmentation of GFAT in individuals with very low thigh fat.

In summary, the segmentation pipeline demonstrated strong performance across cohorts and imaging protocols. VAT, SAT, musculature, and liver were segmented accurately in nearly all cases. Minor segmentation errors were confined to individuals with extreme body composition phenotypes, either due to limited MRI field-of-view in high-BMI individuals or model underperformance in underrepresented anatomical configurations. These findings confirm that our extended segmentation pipeline is robust and suitable for large-scale quantitative body composition analysis.

## Supplementary Tables

**Supplementary Table S1:** Additional descriptive statistics of NAKO participants.

| **Dataset** | **NAKO** |  |
| --- | --- | --- |
| **Sex** | **Male**<br>**(*n* = 14,969)** | **Female**<br>**(*n* = 11,908)** |
| Ethnicity |  |  |
| White | 14,148 (94.52%) | 11050 (92.79%) |
| African | 59 (0.39%) | 34 (0.29%) |
| Indian | 24 (0.16%) | 10 (0.08%) |
| Southeast Asian | 37 (0.25%) | 45 (0.38%) |
| Northeast Asian | 12 (0.08%) | 15 (0.13%) |
| Japanese | 4 (0.03%) | 5 (0.04%) |
| South- & Middle American | 22 (0.15%) | 19 (0.16%) |
| Not specified | 663 (4.42%) | 730 (6.13%) |

**Supplementary Table S2:** Additional descriptive statistics of UKB participants.

| **Dataset** | **UKB** |  |
| --- | --- | --- |
| **Sex** | **Male**<br>**(*n* = 9,094)** | **Female**<br>**(*n* = 9,880)** |
| Ethnicity |  |  |
| White | 8,779 (96.5%) | 9601 (97.2%) |
| Black | 49 (0.5%) | 56 (0.6%) |
| South Asian | 127 (1.4%) | 62 (0.6%) |
| East Asian | 25 (0.3%) | 38 (0.4%) |
| Other | 114 (1.3%) | 123 (1.2%) |

**Supplementary Table S3:** Pearson correlation coefficients between body composition parameters.

| Parameter 1 | Parameter 2 | Male | Female |
| --- | --- | --- | --- |
| ASAT adj. | GFAT adj. | 0.565 [0.556, 0.573] | 0.049 [0.036, 0.063] |
| ASAT adj. | VAT adj. | 0.098 [0.085, 0.110] | 0.275 [0.263, 0.287] |
| ASAT adj. | Muscle adj. | -0.398 [-0.409, -0.388] | -0.132 [-0.145, -0.119] |
| ASAT adj. | LFF adj. | 0.040 [0.027, 0.052] | 0.114 [0.101, 0.127] |
| GFAT adj. | VAT adj. | -0.103 [-0.116, -0.091] | -0.337 [-0.349, -0.325] |
| GFAT adj. | Muscle adj. | -0.407 [-0.418, -0.397] | -0.376 [-0.388, -0.365] |
| GFAT adj. | LFF adj. | -0.088 [-0.100, -0.075] | -0.233 [-0.245, -0.22] |
| VAT adj. | Muscle adj. | 0.115 [0.102, 0.127] | 0.257 [0.245, 0.27] |
| VAT adj. | LFF adj. | 0.350 [0.339, 0.361] | 0.427 [0.416, 0.438] |
| Muscle adj. | LFF adj. | 0.048 [0.035, 0.061] | 0.119 [0.106, 0.132] |

*Note: Values represent Pearson correlation coefficients and 95% confidence intervals between adjusted Z-scores of body composition parameters. ASAT = abdominal subcutaneous adipose tissue; GFAT = gluteofemoral adipose tissue; VAT = visceral adipose tissue; LFF = liver fat fraction; adj. = adjusted.*

**Supplementary Table S4.**
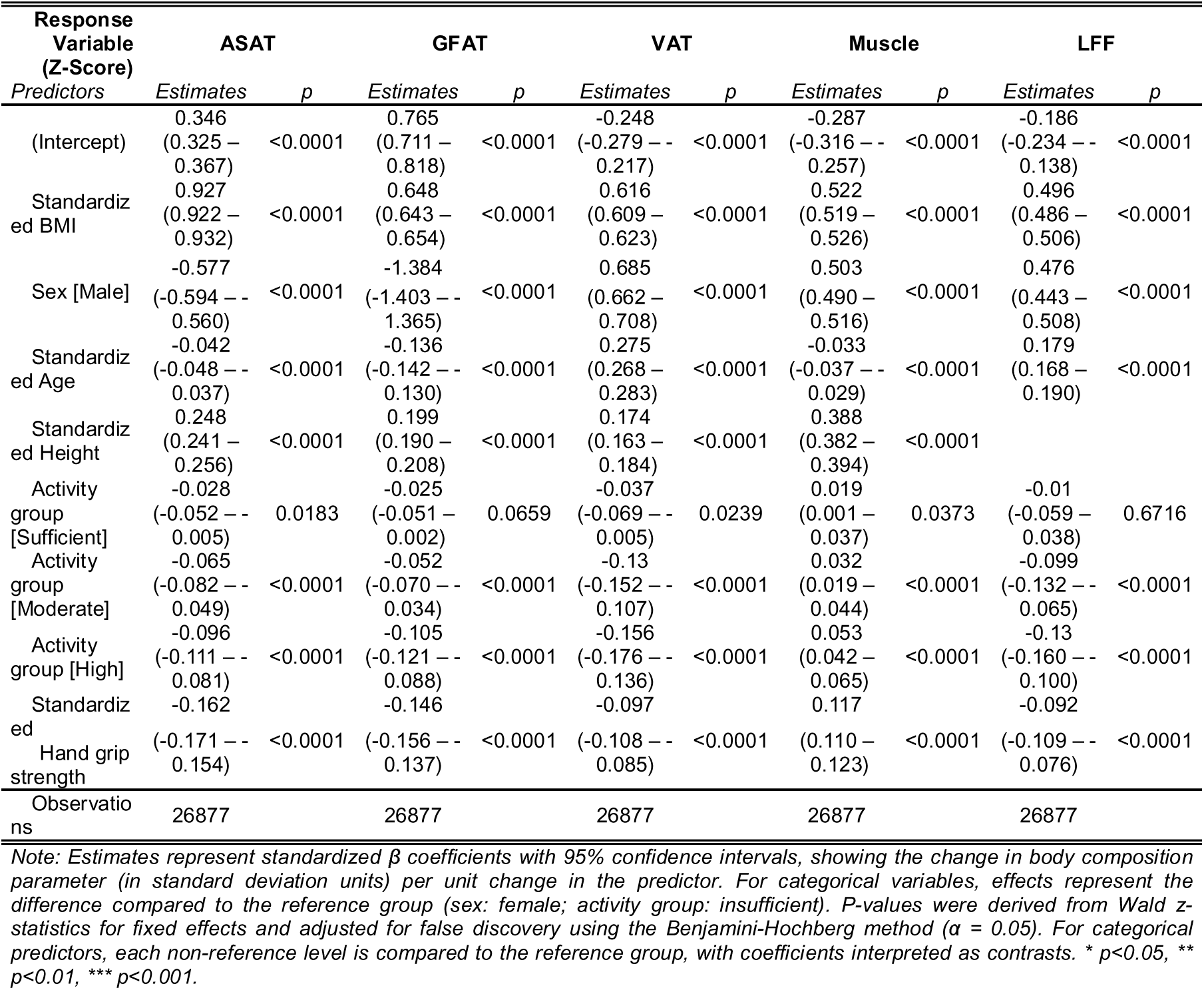
NAKO Linear Mixed-Effects Model Coefficient Estimates.

**Supplementary Table S5.**
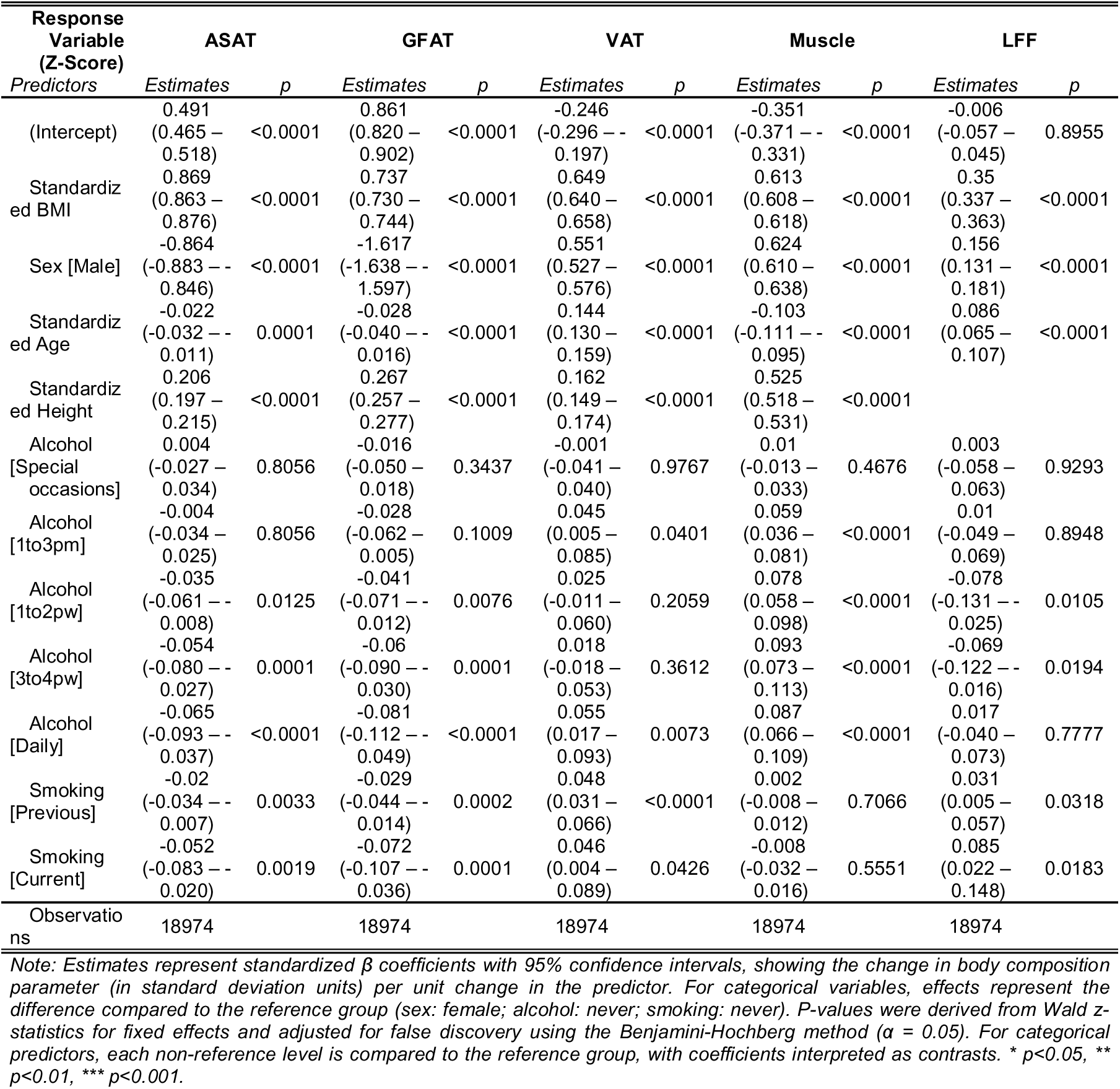
UKB Linear Mixed-Effects Model Coefficient Estimates.

**Supplementary Table S6.**
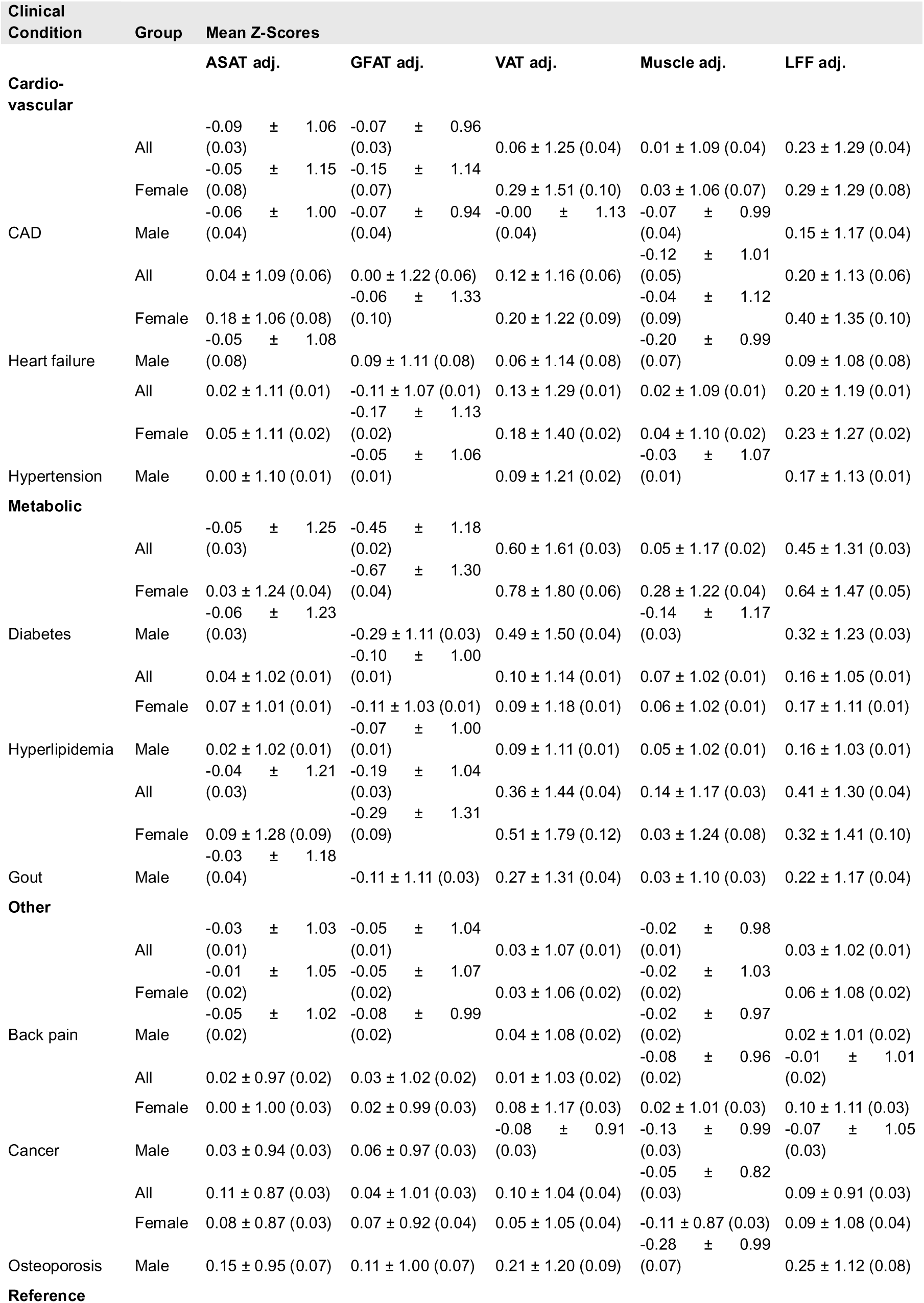
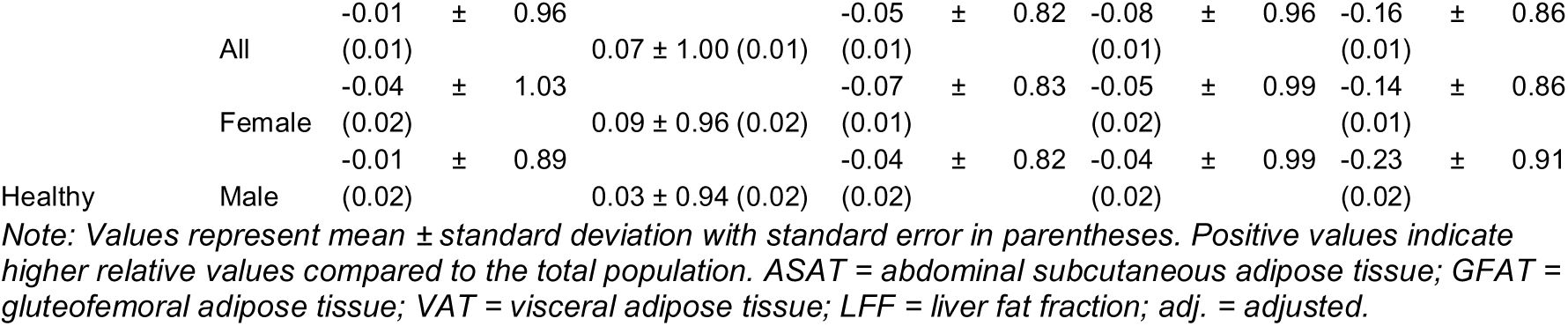
Adjusted BCP Z-score averages for participants with different clinical conditions stratified by sex.

## Supplementary Figures

**Figure S1.**
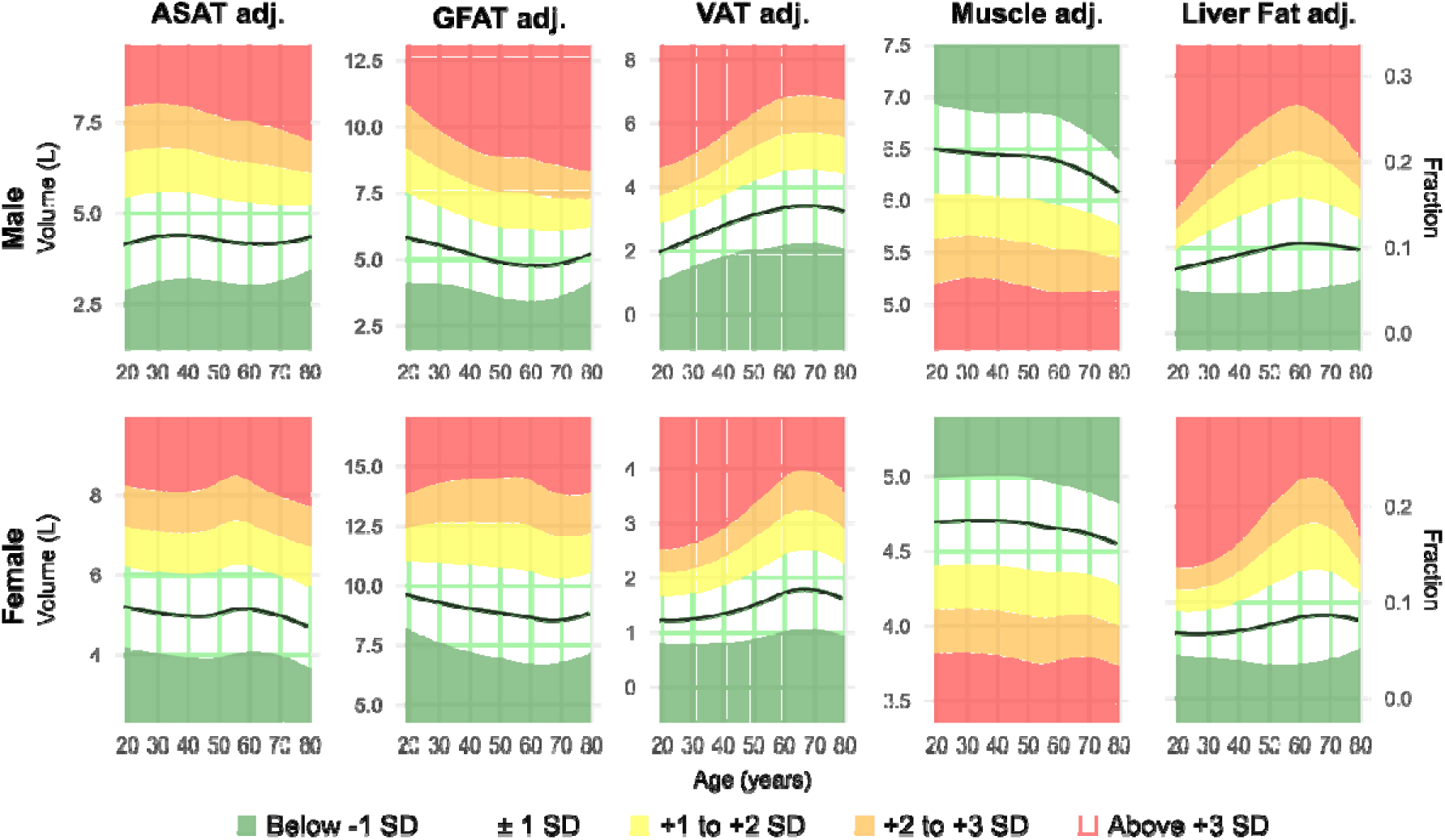
Age-related trends in BMI-adjusted body composition parameters. LOESS curves display the mean and ±1 SD for height-adjusted (except for LFF) and BMI-adjusted body composition parameters across age. Black lines indicate the mean, and shaded areas represent ±1 SD.

**Figure S2.**
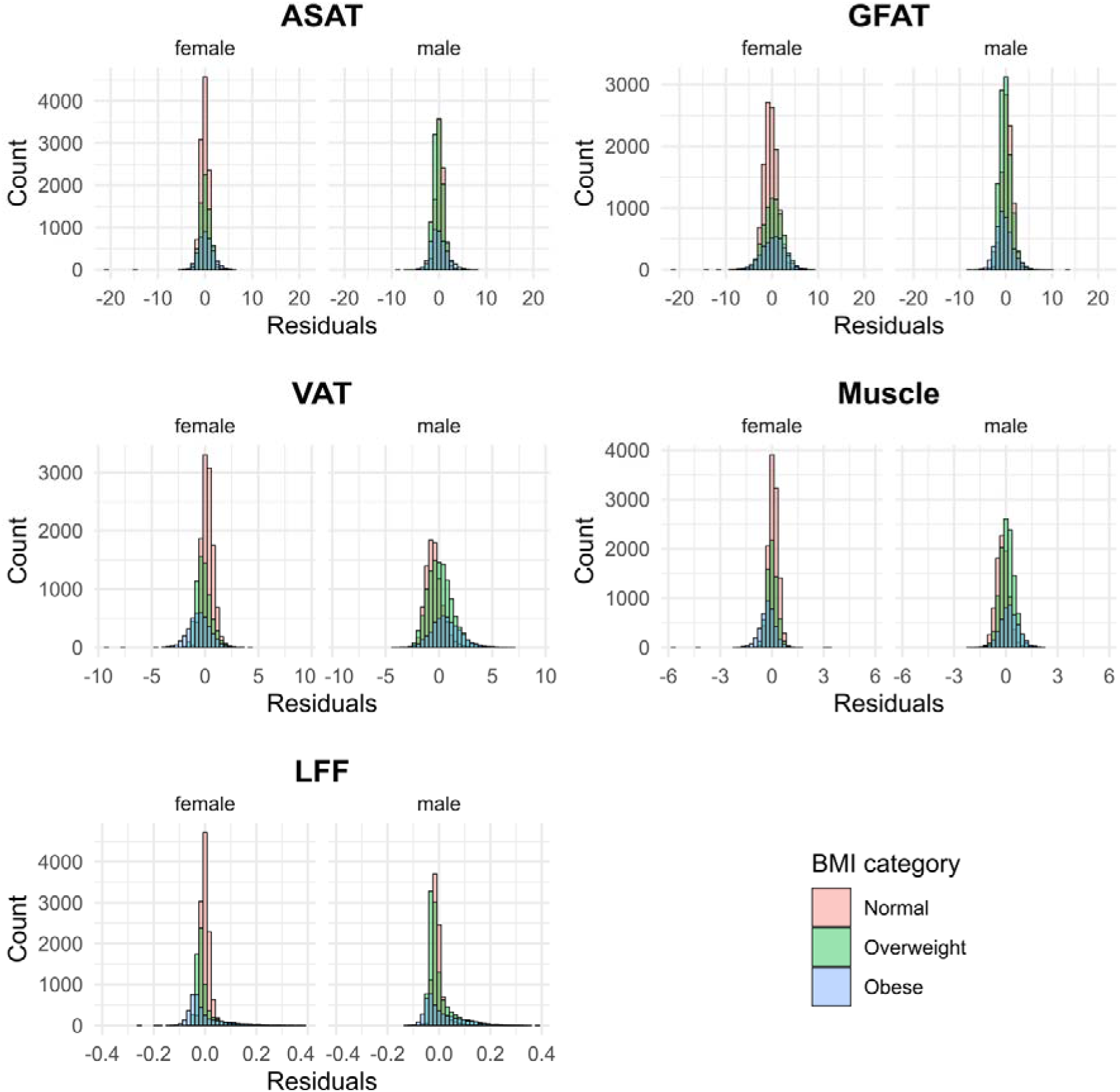
Distribution of body composition parameter residuals after adjustment for anthropometric covariates. Histograms illustrate the distribution of model residuals for abdominal subcutaneous adipose tissue (ASAT), gluteofemoral adipose tissue (GFAT), visceral adipose tissue (VAT), trunk muscle volume, and liver fat fraction (LFF), stratified by sex and BMI category. All measures show approximately normal distributions with minimal systematic bias across BMI categories, validating our statistical approach for isolating depot-specific adiposity measures independent of overall body size.

**Figure S3.**
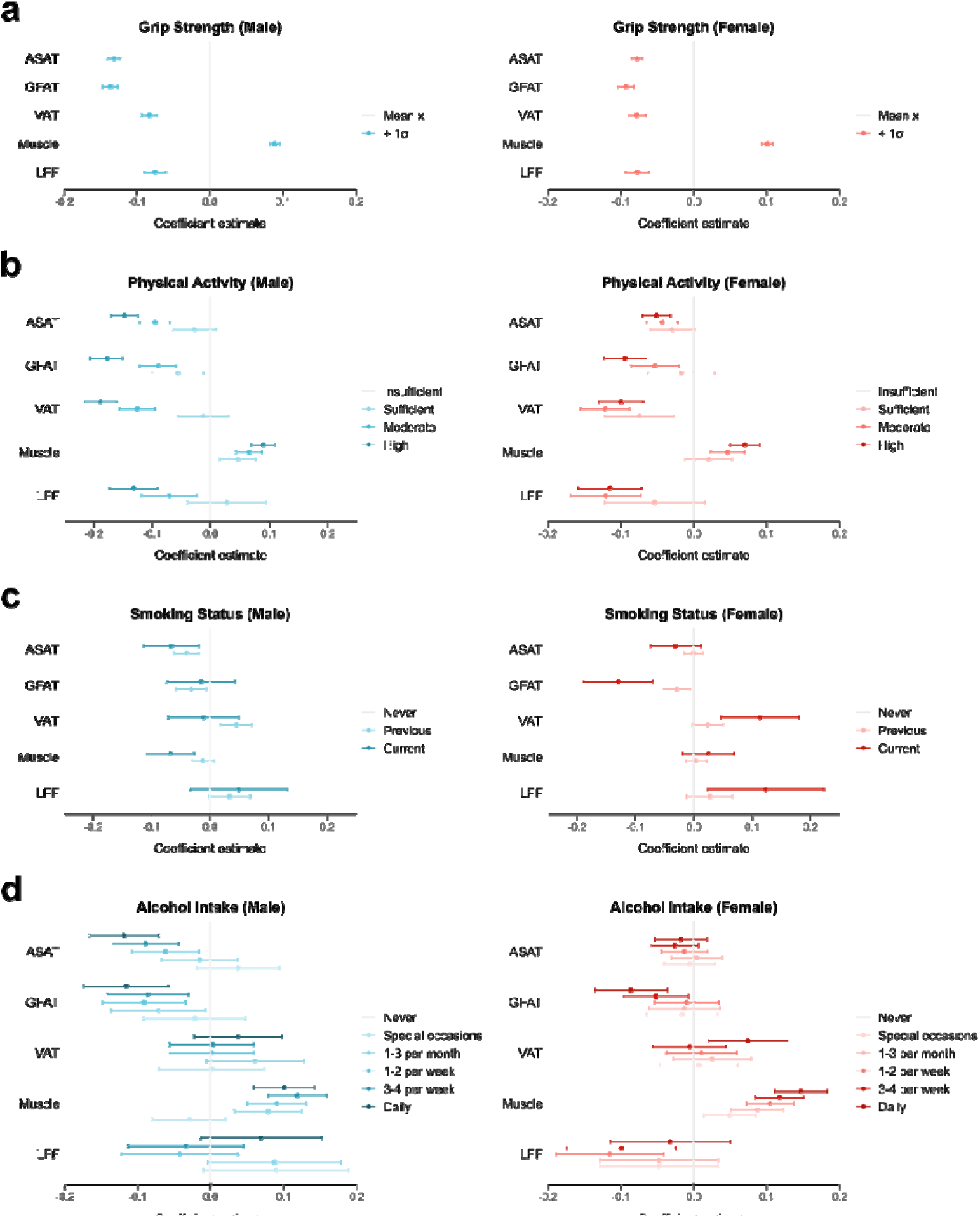
Association of health behaviour indicators with body composition in men and women. Sex-stratified relationships between key health behaviour indicators and MRI-derived body composition parameters based on cohort-specific linear mixed-effects models adjusted for age, height, BMI, and available behavioural factors (maximum hand grip strength & physical activity group (based on MET min. per week of < 600, 600-999, 1000-2999, ≥ 3000) for NAKO; smoking status & alcohol intake frequency for UKB). Imaging centre was included as a random effect. (**a**) Grip strength was inversely associated with all adipose compartments (ASAT, GFAT, VAT, LFF) but positively associated with trunk muscle volume in both sexes. The negative associations with ASAT and GFAT were slightly stronger in men compared to women. (**b**) Dose-dependent relationships between physical activity and body composition, with increasing physical activity levels progressively linked to reduced visceral adiposity and enhanced trunk muscle volume. Interestingly, while men in the highest activity group exhibited even lower amounts of VAT and LFF, albeit with diminishing returns, women did not seem to benefit from further reductions in these adipose tissue compartments with higher activity. (**c**) Smoking was associated with slightly reduced amounts of ASAT and muscle in men. In women, it was linked to decreased amounts of GFAT and notably, increased amounts of VAT and LFF. (**d**) Higher frequency of alcohol intake was associated with reduced amounts of subcutaneous adipose tissue compartments and higher trunk muscle volume in men and women. Associations with VAT and LFF were inconsistent and not statistically robust, with wide confidence intervals suggesting limited precision.

**Figure S4.**
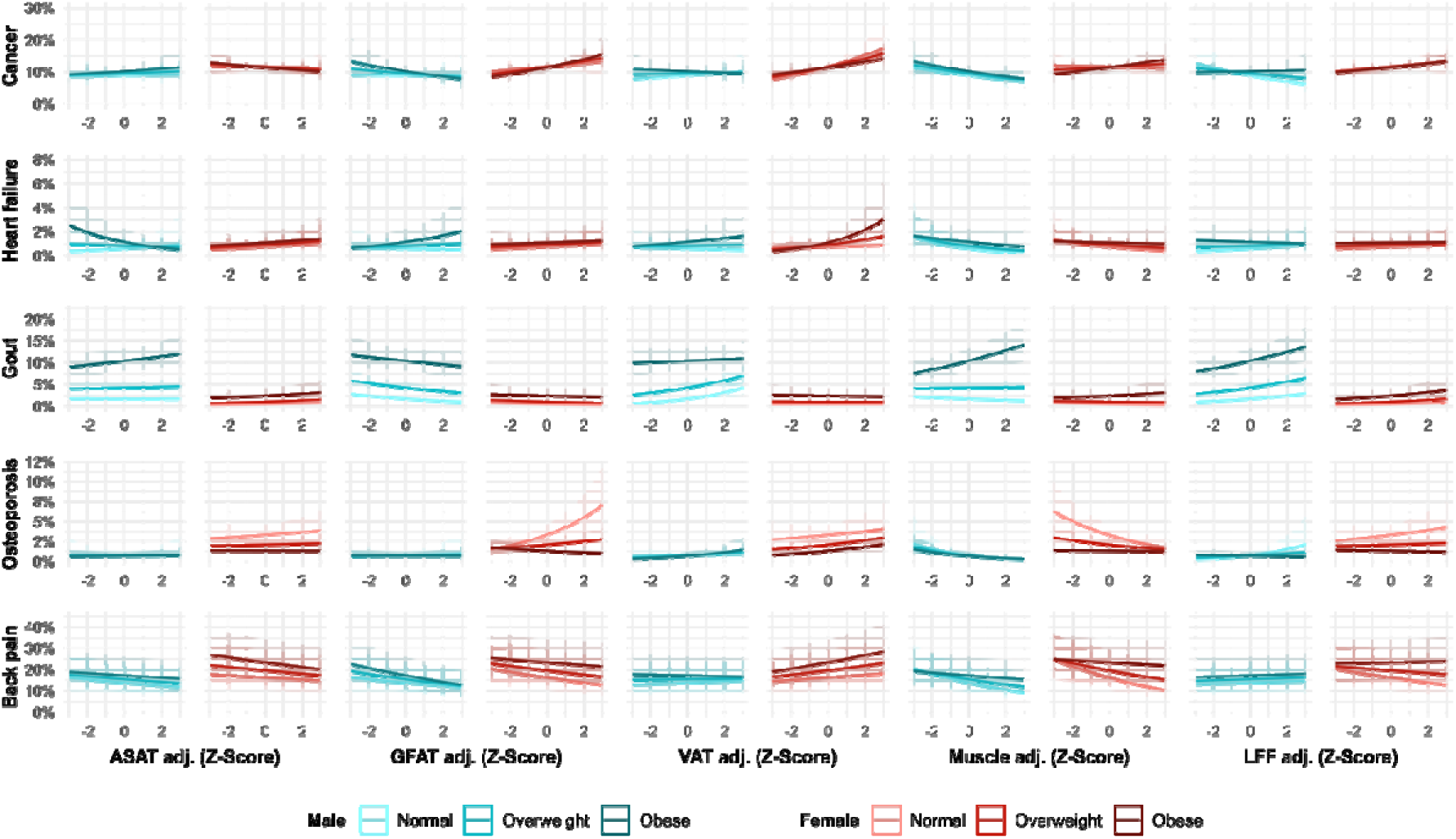
Predicted prevalence plots of other clinical conditions. Plots show the predicted prevalence with confidence intervals of the remaining available clinical conditions (cancer, heart failure, gout, osteoporosis, and back pain) across Z-scores of adjusted body composition parameters (BCPs) based on generalized mixed-effects logistic regression models adjusted for age, BMI, adjusted BCPs, and interaction terms between BMI and each of the BCPs, with imaging centre as a random effect. Data are stratified by sex and BMI category (set at representative values of 20 kg/m^2^ (normal), 27.5 kg/m^2^ (overweight), and 35 kg/m^2^ (obese)).

**Figure S5.**
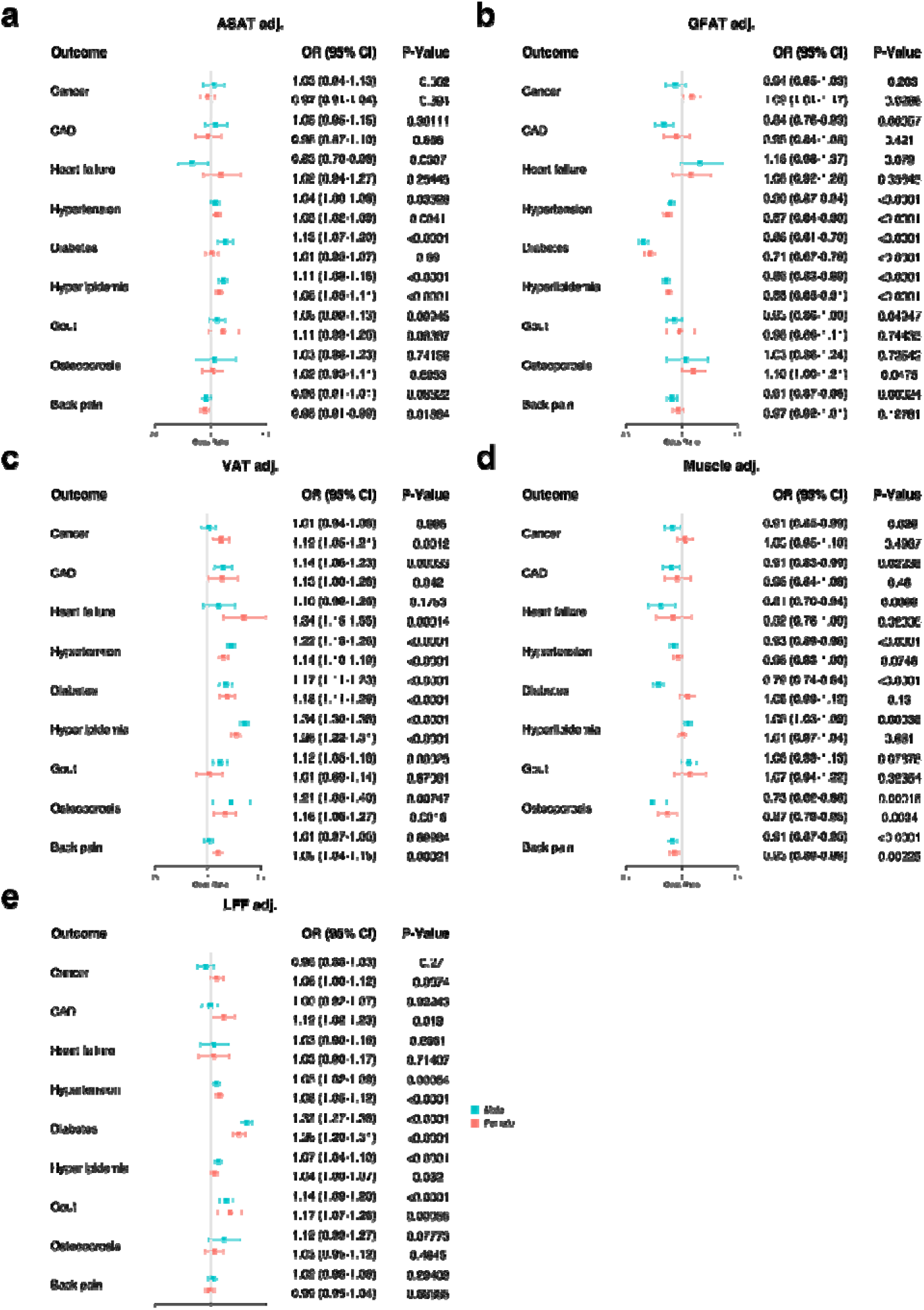
Odds ratios based on adjusted body composition parameters stratified by sex. Forest plots showing associations between adjusted body composition parameters and clinical conditions, stratified by sex (males in blue, females in red). (**a**) ASAT-adj shows modest positive associations with hyperlipidaemia and diabetes, particularly in males. (**b**) GFAT-adj demonstrates inverse effects against metabolic conditions, significantly reducing odds of diabetes (OR=0.66-0.71, p<0.0001) and hyperlipidaemia (OR=0.86-0.88, p<0.0001). (**c**) VAT-adj exhibits the strongest adverse relationships, significantly increasing odds for cardiovascular and metabolic conditions in both sexes, with a notable female-specific association with cancer (OR=1.12, p=0.0012). (**d**) Adjusted trunk muscle volume showed more pronounced effects in men, where it protected against heart failure (OR=0.81, p=0.0068), diabetes (OR=0.79, p<0.0001), and osteoporosis (OR=0.73, p<0.001). (**e**) LFF-adj associates positively with metabolic conditions, particularly diabetes (OR=1.26-1.32, p<0.0001). Results demonstrate distinct sex-specific patterns in how regional adiposity and trunk muscle volume relate to prevalence of cardiometabolic, cancer, and related clinical conditions independent of BMI.

**Figure S6.**
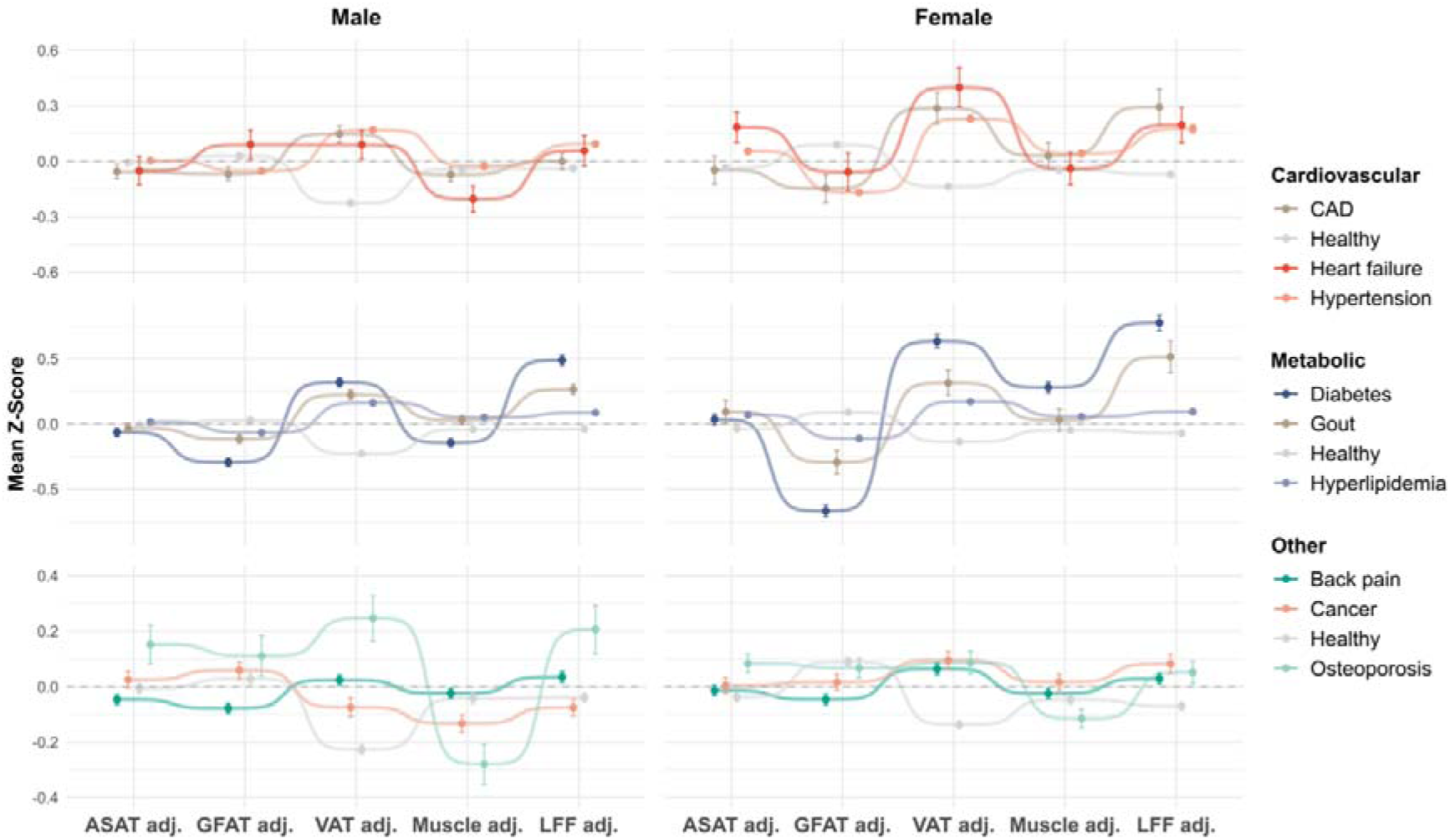
Sex separate body composition phenotypes based on clinical condition status. Body composition phenotypes are presented as mean Z-scores ± standard error of the adjusted body composition parameters (BCPs). The top panel shows cardiovascular conditions (CAD, heart failure, hypertension) compared to healthy controls, revealing distinct patterns particularly in VAT-adj and LFF-adj (elevated) as well as GFAT-adj in women (reduced). The middle panel displays metabolic disorders (diabetes, gout, hyperlipidaemia), with diabetes showing the most pronounced profile characterized by elevated VAT-adj and LFF-adj coupled with substantially reduced GFAT-adj in both sexes, but with greater GFAT deficit in females. The bottom panel presents other conditions (back pain, cancer, osteoporosis) with more subtle deviations from the healthy reference. These condition-specific body composition phenotypes hint at distinct pathophysiological mechanisms underlying different conditions and reveal important sex differences in how adipose distribution relates manifestation of clinical conditions.

**Figure S7.**
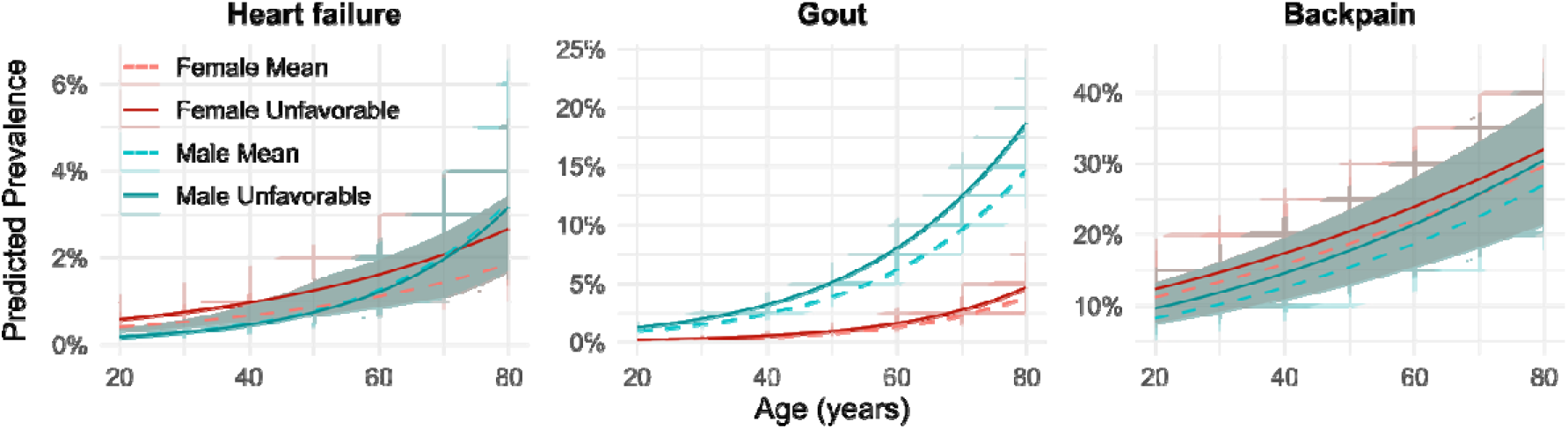
Predicted prevalence plots of other clinical conditions with mean and unfavourable body composition profile stratified by age. Plots show the age-dependent predicted prevalence with confidence intervals of heart failure, gout, and back pain, stratified by sex and body composition profile. Unfavourable profile was defined as +1 SD of adjusted ASAT, VAT, and LFF, with -1 SD of adjusted GFAT and trunk muscle volume. Shaded regions represent 95% confidence intervals. For heart failure, prevalence increased with age in both sexes, with unfavourable body composition associated with higher observed prevalence in women. Gout shows prominent sex dimorphism with substantially higher prevalence in males, particularly those with unfavourable body composition, reaching ∼18% by age 80 compared to ∼5% in females with unfavourable phenotypes. Back pain demonstrates the highest overall prevalence among these conditions, reaching approximately 30% at advanced ages, with age-related increases in both sexes and only a modest increase in predicted prevalence due to unfavourable body composition.

**Figure S8.**
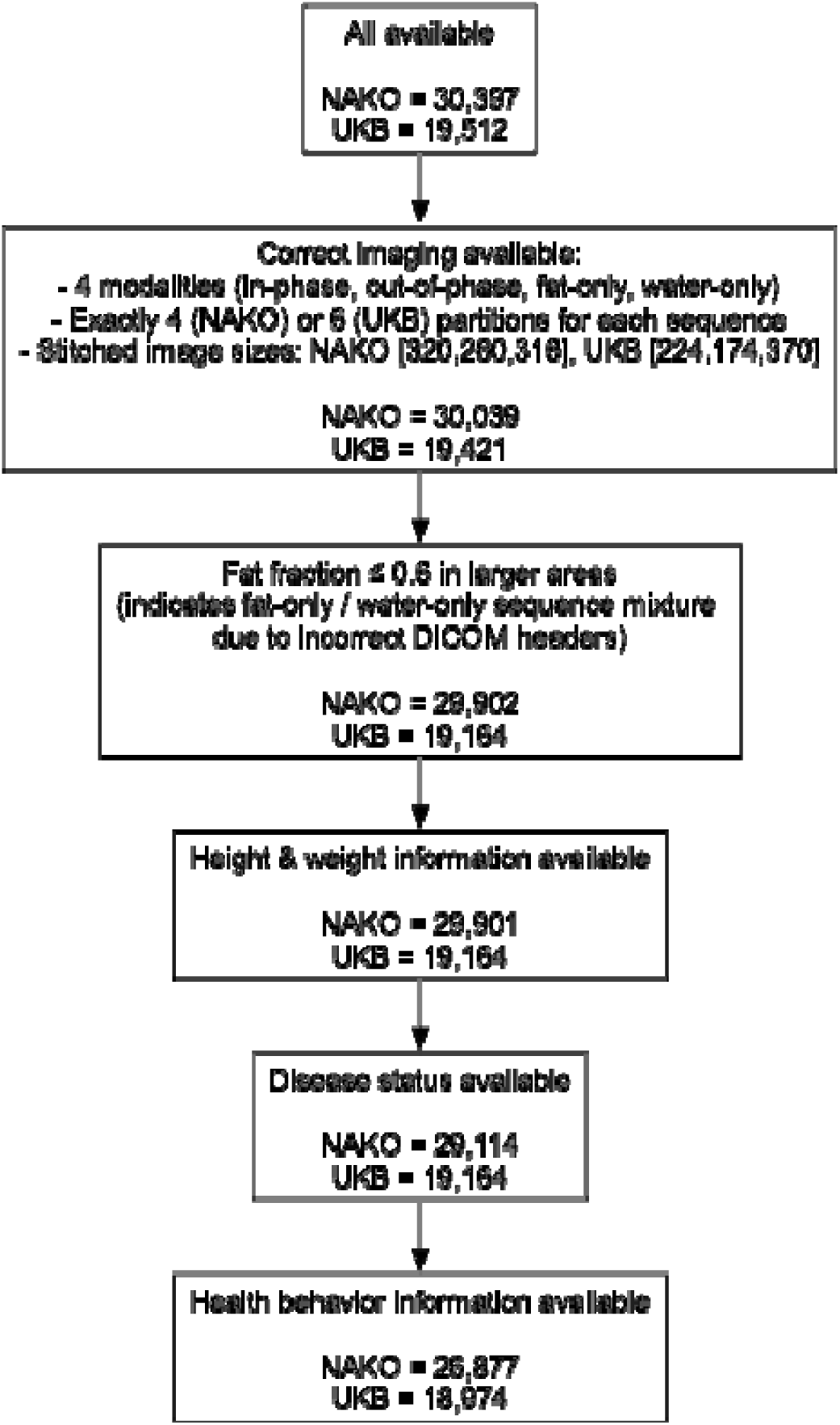
Participant inclusion flowchart. From an initial sample of 30,397 NAKO and 19,512 UKB MRI scans, participants were sequentially filtered: after confirming required imaging modalities and image sizes (yielding 30,039 NAKO, 19,421 UKB), excluding scans with fat fraction ≥ 0.6 in larger areas (29,902 NAKO, 19,164 UKB), and retaining those with height, weight, and clinical condition status data (29,114 NAKO, 19,164 UKB), the final analytic cohorts comprised 26,877 NAKO and 18,974 UKB participants with available health behaviour information.

**Figure S9.**
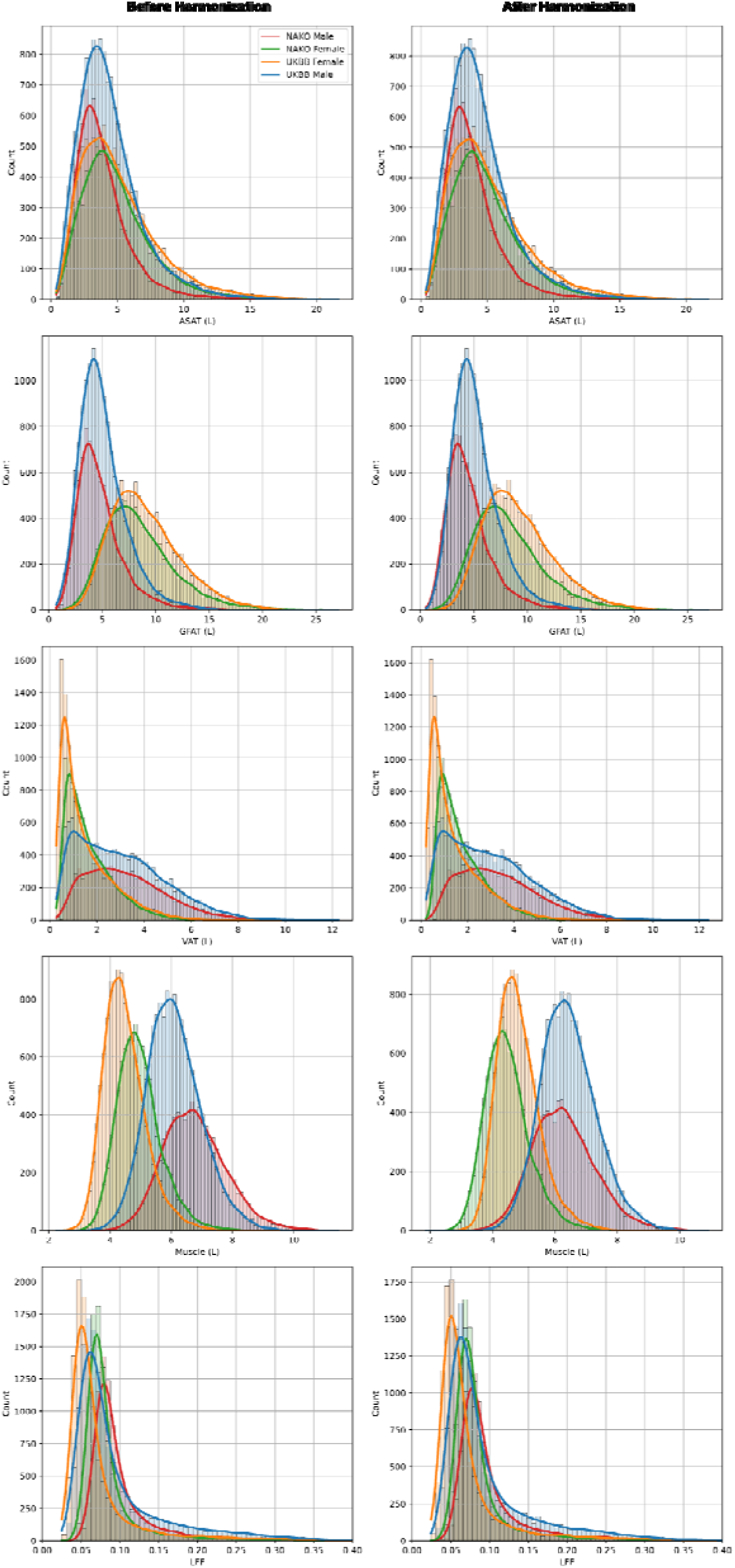
Histograms illustrate the distributions of ASAT, GFAT, VAT, trunk muscle volume, and LFF from NAKO and UKB cohorts, stratified by dataset and sex, before (left panels) and after (right panels) applying ComBat harmonization. This adjustment minimizes site-specific differences while preserving biological variation.

**Figure S10.**
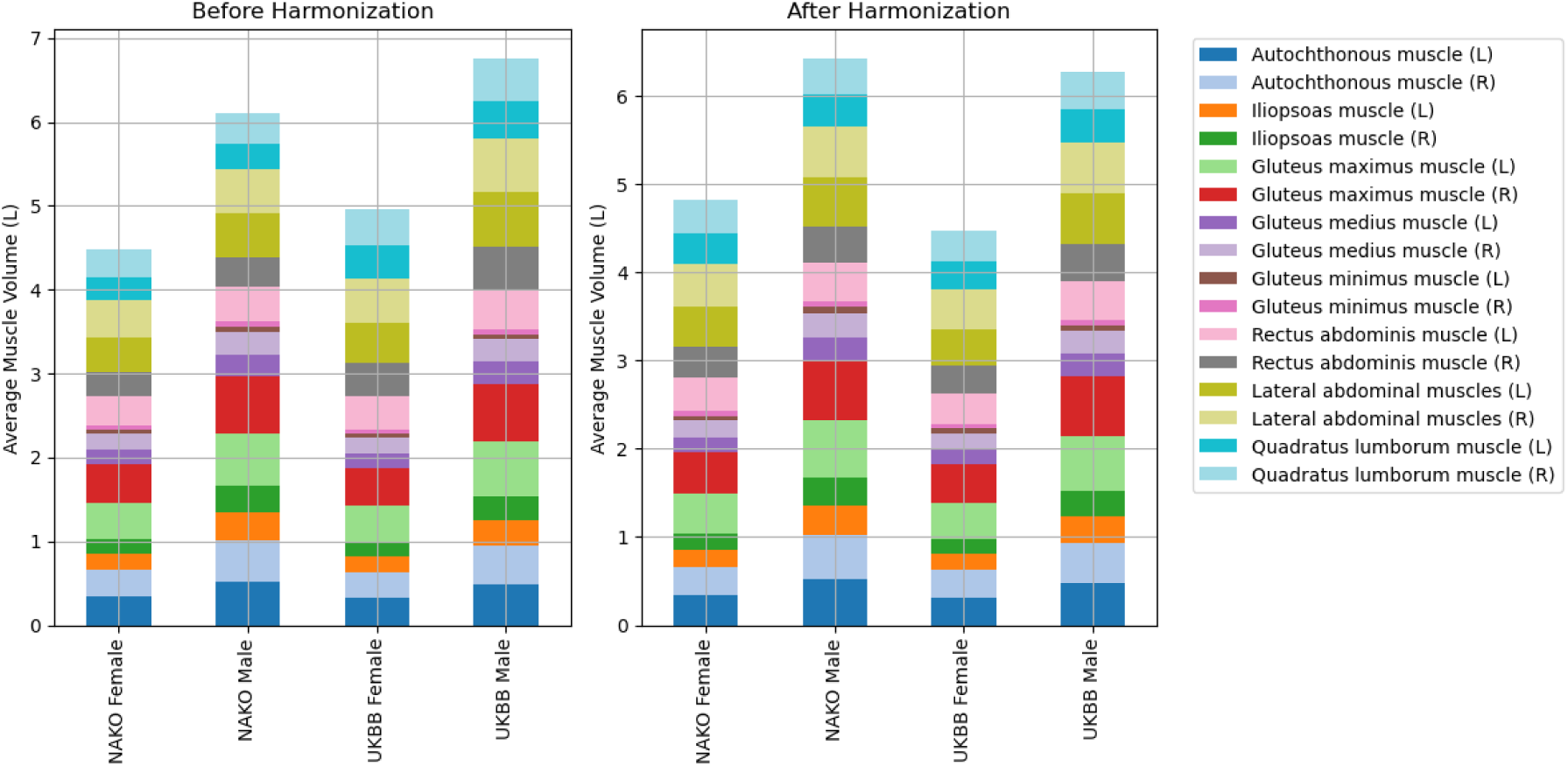
Individual trunk muscle volume components before and after ComBat harmonization. Charts display the contributions of individual muscles form the core, hip, and lower back region to total trunk muscle volume, stratified by dataset and sex. The left panels show the raw values, while the right panels display the data after ComBat harmonization.

